# Inherited genetic risk factors in young-onset lung cancer

**DOI:** 10.64898/2026.04.14.26350822

**Authors:** Myvizhi Esai Selvan, Bonnie Elyssa Gould Rothberg, Abhijit A. Patel, Jian Sang, Amir Horowitz, David C. Christiani, Robert J. Klein, Zeynep H. Gümüş

## Abstract

**Introduction:** Lung cancer is rare before age 45, and its inherited genetic basis remains poorly defined.

**Methods:** We performed whole-genome sequencing in 171 predominantly young-onset lung cancer patients and integrated these data with whole-exome sequencing from six major lung cancer consortia, yielding 9,065 patients. After quality control, analyses focused on 6,545 individuals of European ancestry, the largest ancestral group. We compared the prevalence of rare pathogenic and likely pathogenic (P/LP) germline variants between 186 young-onset (age <45 years) and 6,359 older patients at gene and gene-set levels using Fisher’s exact test, stratified by histology, sex, and smoking status. Polygenic risk scores (PRS) derived from common variants were also evaluated.

**Results:** Young-onset patients carried a higher burden of rare germline P/LP variants in DNA damage response (DDR) genes (including *BRIP1*, *ERCC6*, *MSH5),* and in cilia-related genes, notably *GPR161*. At the pathway level, DDR genes were significantly enriched (OR=1.66, *p*=0.007), with the strongest signal in the Fanconi Anemia pathway and among females (OR=1.96, *p*=0.01). Enrichment was also observed in inborn errors of immunity pathways, with strongest signals in antibody deficiency and the complement system genes. Young-onset patients additionally exhibited higher lung cancer PRS.

**Conclusion:** Young-onset lung cancer exhibits a distinct germline genetic architecture, characterized by enrichment of rare P/LP variants in DDR, cilia-related, and immune pathways, and an elevated lung cancer PRS. These findings support a greater role for inherited susceptibility in early-onset disease and have implications for risk stratification, earlier screening, and precision prevention.

## Introduction

Lung cancer is the leading cause of cancer-related deaths in the United States^1^. While prognosis is substantially better for early-stage disease, most patients are unfortunately diagnosed at advanced stages, when treatment options are limited. In addition to well-established risk factors such as tobacco smoking and environmental or occupational exposures^2^, numerous studies have demonstrated that inherited genetic lesions contribute to lung cancer risk^3–5^. Individuals who carry rare germline pathogenic or likely pathogenic variants (P/LP) in *Fanconi Anemia* genes are at increased risk for squamous cell lung cancer^6^, while those with P/LP variants in *ATM* are at elevated risk for lung adenocarcinoma^7^. Furthermore, we and others have shown that carriers of such variants tend to be diagnosed at younger ages, with an inverse relationship between the number of rare P/LP variants and age at diagnosis^8^. While lung cancer incidence increases strongly with age^9^, its incidence among young adults is rising^10^. Despite this trend, relatively few studies have investigated the germline genetic basis of lung cancer in young adults (defined here as age < 45 years).

Although limited, existing evidence suggests that germline genetic factors may play a distinct role in the development of lung cancer in young adults. First, lung cancer is rare in younger individuals, but relatively common in older adults^11–16^. In the United States, the incidence rate among individuals under 50 years old is 2.2 per 100,000 (CI 2.1-2.2), whereas older age is a recognized risk factor, with a median age of diagnosis of 71 years (https://seer.cancer.gov). Twin studies further support a stronger inherited component in younger patients, reporting greater concordance among young monozygotic twins compared to dizygotic twins, particularly among females^17^. Notably, this difference was not observed among twins older than 50 years^18^. Second, tumors arising in young adults display distinct biological features, including higher frequencies of targetable somatic alterations, such as *EGFR*, *ALK* and *ROS1*, suggesting that underlying germline differences may also exist^19^. Third, young adult lung cancer patients exhibit distinct clinical characteristics: they are more often female, more likely to develop adenocarcinoma and frequently present with advanced-stage disease at diagnosis ^12,20–26^. However, it remains unclear whether delayed diagnosis reflects a lower perceived risk or more aggressive tumor biology with shorter latency.

Together, these observations suggest that lung cancer arising in young adults may involve a stronger and distinct inherited genetic component than disease diagnosed later in life. We hypothesized that young-onset lung cancer reflects an increased burden of inherited susceptibility, characterized by enrichment of rare P/LP variants in certain cancer, immune and developmental pathways, as well as elevated polygenic risk.

To address this, we analyzed germline whole exome (WES) and whole genome sequencing (WGS) data from 9,065 lung cancer patients. This included WGS of germline DNA from 171 young-enriched lung cancer patients, together with WES data from major consortia. We compared the prevalence of rare germline P/LP variants across cancer related genes and functionally related gene-sets between young-onset and older patients, while accounting for sex, smoking status and histology. In addition, we evaluated inherited susceptibility using polygenic risk scores derived from common variants. Together, these analyses provide insights into the germline genetic architecture of lung cancer in young adults, and support future efforts to improve risk stratification, screening, and prevention strategies for individuals at highest risk.

## Materials and Methods

### Study Design

An overview of the study design is shown in Figure 1A, with the analytical workflow summarized in Figure 1B. Briefly, we first performed WGS in the Sinai-Harvard-Yale Young Adult Lung Cancer cohort, comprising 171 lung cancer patients, the majority diagnosed before age 45 years. After sample-level quality control (QC), germline variant calls from this cohort were integrated with WES data from lung cancer cohorts assembled by major international consortia (Supplementary Table S1), resulting in a combined dataset of 9,065 lung cancer patients. Variants were annotated and filtered to retain rare P/LP variants based on ClinVar annotations^27^. The prevalence of P/LP variants was then compared between young adults (< 45 years) and older patients (≥ 45 years) with lung cancer at gene and gene-set levels. Analyses were stratified by histology, smoking status and sex.

**Figure 1.**
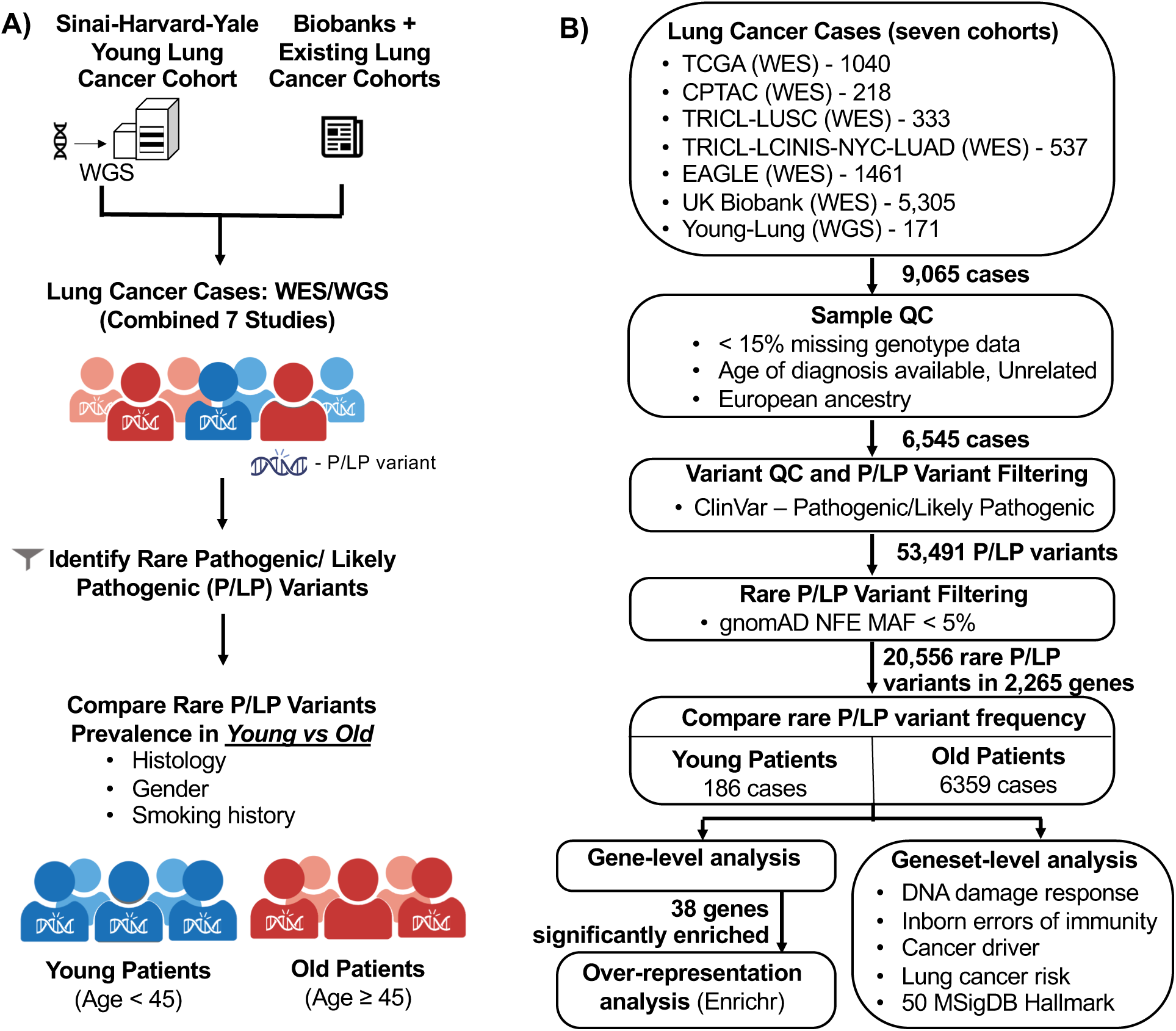
A) Study design B) Study analysis pipeline.

### Sinai-Harvard-Yale Young Adult Lung WGS Cohort Analysis

Germline WGS was performed on 171 samples biobanked at Yale Medical School, Harvard T.H. Chan School of Public Health (Boston Lung Cancer Study-BLCS) and Icahn School of Medicine at Mount Sinai. Yale samples were collected under HIC #1010007459. The BLCS cohort was approved by the Mass General Brigham (MGB) IRB, protocol #1999P004935. Variants were called from raw sequencing data using a standardized analysis pipeline described below.

#### Library Preparation and Whole Genome Sequencing (WGS) Protocol

WGS was performed at the Beijing Genomics Institute (BGI) China, generating 100bp paired-end reads with an average of 90G clean reads per sample. Briefly, samples were first tested for DNA concentration, sample integrity and purity. DNA concentration was detected by a fluorometer or Microplate Reader (e.g. Qubit Fluorometer, Invitrogen). Sample integrity and purity were evaluated using 1% agarose gel electrophoresis (Voltage: 150V, 40 mins). Next, 1μg of genomic DNA per sample was randomly fragmented by Covaris, followed by size selection (200-400bp) with the Agencourt AMPure XP-Medium kit. Fragmented DNA underwent end-repair, 3’ adenylation, adaptor ligation, and PCR amplification. PCR products were purified by the Agencourt AMPure XP-Medium kit, heat denatured, and circularized using a splint oligonucleotide sequence to generate single-stranded circular DNA (ssCir DNA). Libraries passing QC were sequenced on the BGISEQ-500 platform. ssCir DNA molecules were amplified into DNA nanoballs (DNB) containing more than 300 copies through rolling-cycle replication and loaded onto patterned nanoarrays by using high density DNA nanochip technology. Paired-end 100bp reads were obtained by combinatorial Probe-Anchor Synthesis (cPAs).

#### Variant Calling

Raw sequencing files were processed using a standardized WGS pipeline developed at Mount Sinai, following guidelines for functionally equivalent pipelines^28^. Briefly, reads were aligned to reference genome build 38 using BWA-MEM^29^ v0.7.15 with parameters “-K 100000000 -Y” and processed with samblaster “-a –addMateTags”. Duplicate marking and sorting of bam files were performed using GATK 4 MarkDuplicatesSpark^30^. Base quality score recalibration was performed using GATK 4^31,32^, and the resulting recalibrated files were compressed to CRAM format using samtools. For each individual, GVCF files were generated using GATK HaplotypeCaller, followed by joint genotyping with GenotypeGVCFs. Variant quality score calibration was performed separately for SNPs and indels using GATK-recommended parameters and resources, including HapMap 3.3, Omni 2.5, 1000 Genomes, dbSNP 138 for SNPs, and Mills/1000 Genomes, dbSNP and Axiom for indels^31,33^.

#### Sample QC

Consistent with our prior studies on germline sequencing datasets^6–8^, samples with 15% or more missing genotype data were excluded. Duplicates or related individuals up to second degree were removed based on relatedness analysis using KING^34^. To remove any bias that may arise due to population stratification, Principal Component Analysis (PCA) was performed, and analyses were focused on the largest ancestry group, individuals of European ancestry. For PCA analysis, we used our established analysis pipeline^6–8^, using 1000 Genome dataset as reference and common SNPs (minor allele frequency (MAF) > 5%), with smartpca from EIGENSOFT v5.0.1. After QC, 149 individuals (134 LUAD, 14 LUSC, 1 other) with available age-at-diagnosis information were retained (Supplementary Figure S1).

#### Variant QC

For WGS samples that passed QC, variants were filtered to retain high-quality genotype and variant calls. Genotype-level filters included genotype quality ≥20, read depth ≥10, and allelic depth of alternate allele ≥4. Site-level filters included quality score ≥30, quality by depth score ≥2, variant tranche <99%, and missingness < 15%. Additional filters were applied for SNPs (strand odds ratio ≤ 3, Fisher strand ≤ 60, mapping quality ≥40, read position rank sum ≥–8, mapping quality rank sum ≥–12.5) and indels (Fisher strand ≤ 200, read position rank sum ≥–20). For heterozygous genotypes, alternate allele ratios between 0.30 and 0.8 were required.

### Combined Study Cohort Overview

To increase statistical power, germline WGS data from the Sinai-Harvard-Yale Young Adult cohort were combined with germline WES data from six major lung cancer consortia with publicly available germline data (Supplementary Table S1).

1. *The Cancer Genome Atlas (TCGA*) *cohort*^8^ includes 1,040 participants with lung cancer (546 LUAD and 494 LUSC). After sample QC and ancestry filtering, 844 patients of European ancestry were included in the analysis (410 LUAD and 434 LUSC).
2. *Clinical Proteomic Tumor Analysis Consortium (CPTAC) cohort*^35^ includes 218 patients (110 LUAD and 108 LUSC). After sample QC and ancestry filtering, 123 patients of European ancestry were retained (41 LUAD and 82 LUSC).
3. *Transdisciplinary Research Into Cancer of the Lung (TRICL) LUSC cohort*^6^ comprises 333 LUSC participants. After sample QC and ancestry filtering, 318 patients (LUSC) of European ancestry were included in the analysis.
4. *TRICL-LCINIS-NYC LUAD cohort*^7^ includes 537 LUAD cases aggregated from the TRICL database, the Lung Cancer in Northern Israel (LCINIS) study, and three NYC institutions: Memorial-Sloan Kettering Cancer Center, Weill Cornell Medical College and Icahn School of Medicine at Mount Sinai. After sample QC and ancestry filtering, 483 participants of European ancestry were included in the analysis.
5. *Environment and Genetics in Lung cancer Etiology (EAGLE) cohort*^36^ includes 1,461 primary lung cancer cases from Italy, with no evidence of population stratification among cases. For the current analysis, we filtered out large cell carcinoma, small cell carcinoma and those unclassifiable cases of poorly differentiated/ epithelial/ neuroendocrine/ mucoepidermoid/ non-small cell/ pleomorphic/ sarcomatoid 1,341 patients were included (662 LUAD, 479 LUSC and 200 Other mixed/unspecified non-small cell lung carcinoma).
6. *UK Biobank (UKB)*^37^. Of 502,173 UKB participants, 5,305 lung cancer cases were identified in the cancer registry using ICD-10 code C34 (Data Field 40006), and ICD-9 code 162 (Data Field 40013). Cases were then filtered to include only those with: non-small cell carcinoma histology (Data Field 40011; codes 8046, 8070, 8071, 8140, 8250, 8253, 8260, 8310, 8440, 8480, 8550, 8560), malignant primary site behavior (Data Field 40012), Caucasian genetic ethnic grouping or self-reported as British, Irish or any other white background (Data Field 22006), no second degree or closer relatedness, and available WES data. After applying these filters, 3,287 participants were included in our analysis (1,964 LUAD, 926 LUSC and 397 Other).

#### Variant QC of EAGLE and UKB cohorts

For variant QC of the EAGLE cohort, we filtered for variants with read genotype quality ≥20, read depth ≥10, allelic depth of alternate allele ≥4, alternate allele ratio ≥ 0.30 and ≤ 0.8 for heterozygous genotypes; sites with: quality score ≥50, missingness < 15%. For the UKB cohort, to remove variants that failed QC we used the helper file *ukb23158_500k_OQFE.90pct10dp_qc_variants.txt*, and to identify loss of function (LOF) variants, we used *ukb23158_500k_OQFE.annotations.txt.gz*. As part of UKB variant QC, we also excluded variants with read depth < 10, allelic depth of alternate allele < 4 and alternate allele ratio < 0.30 and > 0.8 for heterozygous genotypes.

#### Integration of WGS and WES Data and Variant Annotation

Variants passing sample- and variant-level QC in each cohort were merged and filtered to retain P/LP variants (Supplementary Table S1). A variant was classified as P/LP if it was annotated as pathogenic or likely pathogenic in the ClinVar database^27^, or it was a frameshift or stopgain variant located 5′ of a known pathogenic loss-of-function (LOF) (nonsense and frameshift) variant in ClinVar. ClinVar annotations were obtained using the Annovar^38^ tool (version: clinvar_20240520). To focus on rare variation, variants with minor allele frequency (MAF) ≥ 5% in gnomAD non-Finnish European cohort were excluded (Supplementary Table S2).

### Gene and gene-set level comparisons of rare germline P/LP variants

Patients were stratified by age at diagnosis into young adults (< 45 years) and older patients (age ≥ 45 years). To assess whether inherited susceptibility differs by age of onset, we compared the gene-level prevalence of rare germline P/LP variants between these groups using Fisher’s exact test. Genes with a single P/LP variant site from one cohort in both age groups were excluded to reduce sparse-data bias. This analysis identified genes with significant age-specific enrichment between the two groups (*p* ≤ 0.05; Supplementary Table S3). We then performed over-representation analysis of the significantly enriched genes using Enrichr^39^, testing multiple pathway and ontology databases, including Gene Ontology (GO) Biological Process 2025, GO Cellular Component 2025, GO Molecular Function 2025, Reactome pathways 2024 and Jensen DISEASES Experimental 2025 (*p* ≤ 0.01 and overlapping genes ≥ 2; Supplementary Table S4). To capture more subtle, but functionally consistent age-related differences, we next compared gene-set level prevalence of rare germline P/LP variants between these groups using Fisher’s exact test. Given the well-established role of DNA repair pathways in cancer development, we first examined variants in the DNA Damage Response (DDR) gene set^35^, comprising ∼400 genes curated from 18 pathways involved in DNA damage detection, signaling, and repair. These pathways included homologous recombination (HR), non-homologous end joining (NHEJ), nucleotide excision repair (NER), bases excision repair (BER), mismatch repair (MMR), Fanconi Anemia (FA), translesion synthesis (TLS), direct repair (DR), Microhomology-mediated end joining (MMEJ), alternative end joining (alt-EJ), telomere, topological stress, meiotic recombination, nucleotide metabolism, DDR signaling, DNA replication, core mitotic DDR and others involved in cellular coordination and molecular responses to DNA damage.

We then assessed genes associated with inborn errors of immunity^40,41^ (IEI; ∼550 genes), given the importance of immune surveillance in tumor suppression and the link between germline immune defects and cancer predisposition. Analyses were performed both collectively and within ten distinct phenotypic categories: immunodeficiencies affecting cellular and humoral immunity; combined immunodeficiencies with associated or syndromic features; predominantly antibody deficiencies; diseases of immune dysregulation; congenital defects of phagocyte number, function, or both; defects in intrinsic and innate immunity; autoinflammatory disorders; complement deficiencies; bone marrow failure; and phenocopies of primary immunodeficiencies.

Finally, we evaluated several biologically and clinically relevant gene sets, including cancer drivers, known lung cancer risk genes, and hallmark gene sets. The cancer drivers gene set included 299 genes identified as drivers in a pan-cancer TCGA study^42^. The lung cancer risk gene set included ∼150 genes identified from Genome-Wide Association Studies (GWAS) of lung cancer susceptibility^43,44^. The complete gene lists for these gene-sets are in Supplementary Table S5. To test canonical biological pathways, we evaluated 50 Hallmark gene sets from MSigDB (https://www.gsea-msigdb.org/).

### Polygenic Risk Score (PRS) Calculation

In addition to detailed rare P/LP variants analyses, we assessed differences in common variant genetic risk using polygenic risk scores. Towards this end, we calculated lung cancer PRSs using PGS004860 (1,143,554 variants-PGP000607^45^) from the PGS Catalog. These analyses were restricted to the Sinai-Harvard-Yale Young Adult Lung WGS Cohort, which had the highest proportion of young adult lung cancer patients with available WGS data. An additional 500 European-ancestry individuals from the 1000 Genomes Project, jointly called with the Sinai-Harvard-Yale cohort, were included as non-lung cancer controls. PRSs were calculated using PLINK allelic scoring, and score distributions were compared across three groups: young adults (n=84), older lung cancer patients (n=65), and controls (n=500) using the Mann-Whitney test.

## Results

### Study Design and Cohort Characteristics

To characterize the germline genetic architecture of young-onset lung cancer, we performed WGS on germline DNA from 171 predominantly young lung cancer patients and integrated these with germline variant calls from six major previously published lung cancer sequencing cohorts^6–8,35–37^. This resulted in a combined dataset of 9,065 patients, representing the largest germline sequencing cohort of lung cancer patients assembled to date across age groups. Within this cohort, we investigated the distribution of rare germline pathogenic or likely pathogenic (P/LP) variants between young adult and older patients, stratifying analyses by histology, sex, and smoking status. An overview of the study design is shown in Figure 1A, with the analytical workflow summarized in Figure 1B.

After sample-level QC, our analyses focused on the largest ancestral group, which was 6,545 individuals of European ancestry, including 186 young-onset patients (age <45 years) and 6,359 older patients (age ≥45 years) (Supplementary Table S1). The mean age at diagnosis was 66.7 years (standard deviation = 9.0; range: 23.2–91.0). Overall, 58.2% of patients were male and 41.8% female. The cohort included 3,694 patients with lung adenocarcinoma (LUAD), 2,253 with lung squamous cell carcinoma (LUSC), and 598 with other non-small cell lung cancer subtypes. Smoking status was available for 6,468 patients (3,655 LUAD and 2,221 LUSC), of whom 87.8% were current or former smokers (3,031 LUAD and 2,117 LUSC) and 12.2% were never-smokers. Clinical characteristics are summarized in Table 1.

**Table 1:**
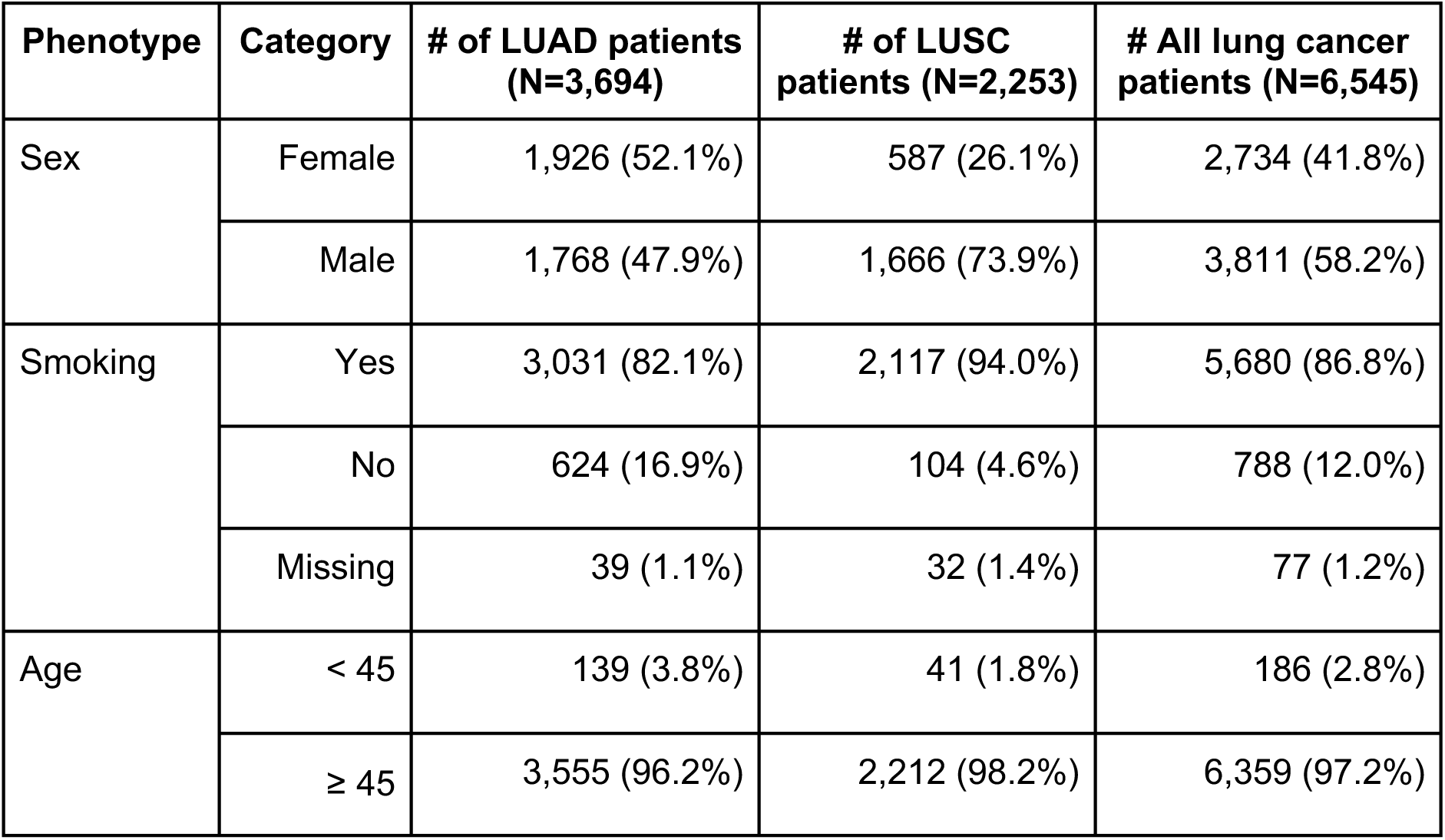
Clinical characteristics of the study cohort.

Across the cohort, we identified 53,491 germline P/LP variants spanning 2,276 genes. Of these, 20,556 variants in 2,265 genes were rare (gnomAD Non-Finnish European (NFE) MAF < 5%), and present in 92.6% of patients (6,062/6,545). This high prevalence reflects the inclusion of ClinVar-annotated variants across unselected genes. At a more stringent threshold (MAF < 1%), 17,559 variants remained, present in 2,254 across 88.8% of patients (5,814/6,545). The distribution of P/LP variants is summarized in Supplementary Table S2. For downstream analyses, we focused on rare P/LP variants with gnomAD NFE MAF < 5%.

### Young-onset lung cancer exhibits distinct patterns of rare germline variation

We next assessed whether the burden of rare P/LP variants differed between young-onset and older lung cancer patients, conducting analyses at both gene- and gene-set levels to capture signals across multiple scales. At the gene level, P/LP variants were identified across 2,265 genes: 34 genes were observed only in young adult patients, 1,898 only in older patients, and 333 genes were observed in both age groups. Among genes observed in both groups, 36 showed a higher prevalence of rare P/LP variants in young adult patients (p<0.05; Figure 2A, Supplementary Table S3A), while two (*HYDIN* and *IVD*) were more prevalent in older patients (Figure 2A, Supplementary Table S3B). Of the genes enriched in young adults, five (*FLG, TPK1, GPR161, PDE11A* and *LFNG*) exceeded 3% prevalence, with *GPR161*, *LFNG* and *TPK1* showing the strongest enrichment (*p* < 1e-4).

**Figure 2.**
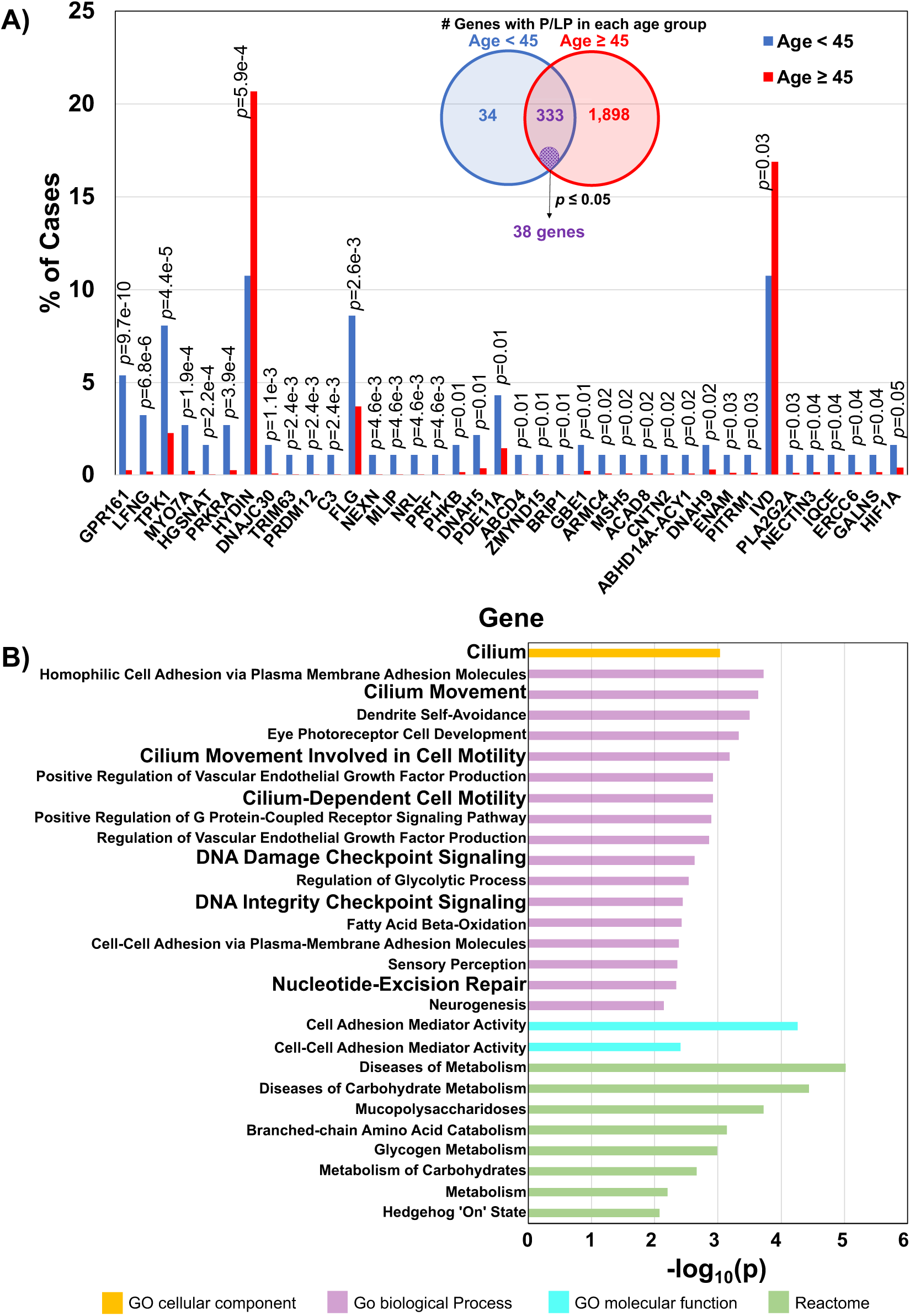
A) Genes with rare P/LP variants in both young adult and older lung cancer patients (Blue: < 45 years; red: ≥ 45 years), highlighting 38 genes with significantly different prevalence (*p* ≤ 0.05). B) Over-representation analysis of the 38 genes using Enrichr.

To better understand the biological processes underlying these observations, we performed over-representation analysis on the 38 genes enriched between the two age groups. This analysis identified enrichment in pathways related to DNA Damage Repair (DDR) and cilia-associated processes (*p* < 0.01; number of genes ≥ 2; Figure 2B; Supplementary Table S4). Notably, multiple genes involved in ciliary structure and function (including *DNAH5*, *GPR161*, *IQCE*, *DNAH9*) were more frequently altered in younger cases. Disease enrichment analyses further linked young-onset enriched genes to lung-related conditions, including lung adenocarcinoma (*MSH5* and *NEXN*) and chronic obstructive pulmonary disease (COPD) or obstructive lung disease (*GALNS*, *DNAH5*, *MSH5*, *IQCE*, *NEXN*, and *CNTN2; Jensen DISEASES* Experimental database), supporting the biological relevance of these findings (Supplementary Table S4).

### Young-onset patients exhibit the strongest enrichment for rare germline P/LP variants in DNA Damage Response gene sets

Beyond over-representation in individually significant genes, we next performed pathway enrichment across all genes with P/LP variants (see Methods, Supplementary Table S5) across the full cohort. Stratification by age group revealed that the strongest and most consistent enrichment in young-onset patients occurred in the DDR gene set (Figure 3, Supplementary Table S6). Across all lung cancers, younger age at diagnosis was associated with increased prevalence of DDR P/LP variants (OR = 1.66; *p*-value = 0.0070; 95% CI: 1.13 - 2.40) (Figure 4A). Notably, this enrichment was observed across multiple stratifications, including histology, sex and smoking status, supporting robustness of the signal. The effect was particularly pronounced in LUSC (OR=3.24; *p*-value=0.0009; 95% CI= 1.55 - 6.48), while a similar but non-significant trend was observed in LUAD (OR = 1.34; *p*-value = 0.21; 95% CI: 0.81 - 2.11) (Figure 4A). Consistent with this pattern, age-stratified analyses revealed an age-dependent gradient, with progressively decreasing prevalence of DDR P/LP variants with increasing age. The most significant difference was observed between the extreme age groups (age <45 vs ≥65 years) (Supplementary Figure S2).

**Figure 3.**
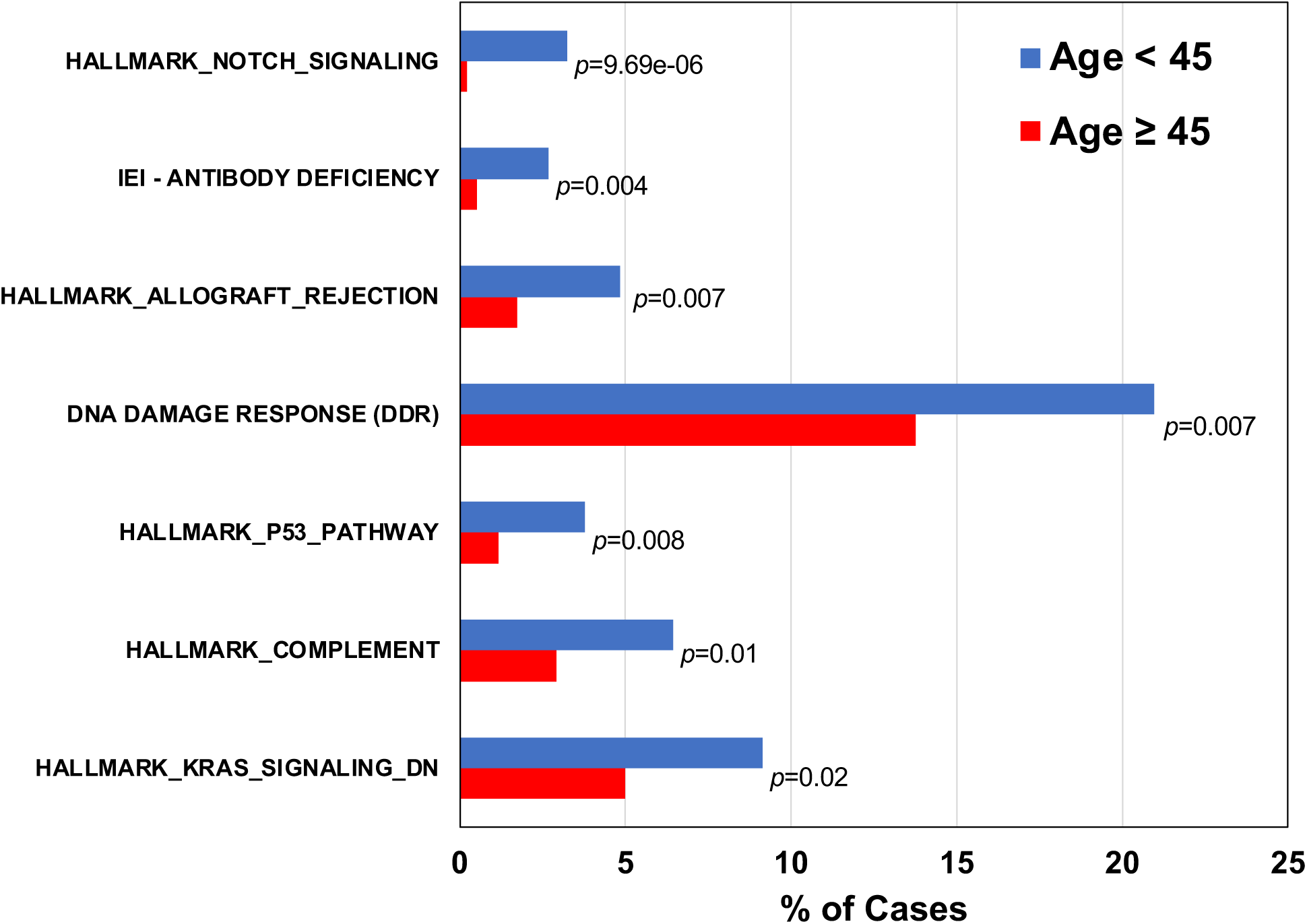
Gene-set level distribution of rare P/LP variants in lung cancer patients. Blue: < 45 years; red: ≥ 45 years. Listing gene-sets that show a statistically significant difference in prevalence (*p* ≤ 0.05) between young adults and older lung cancer patients.

**Figure 4.**
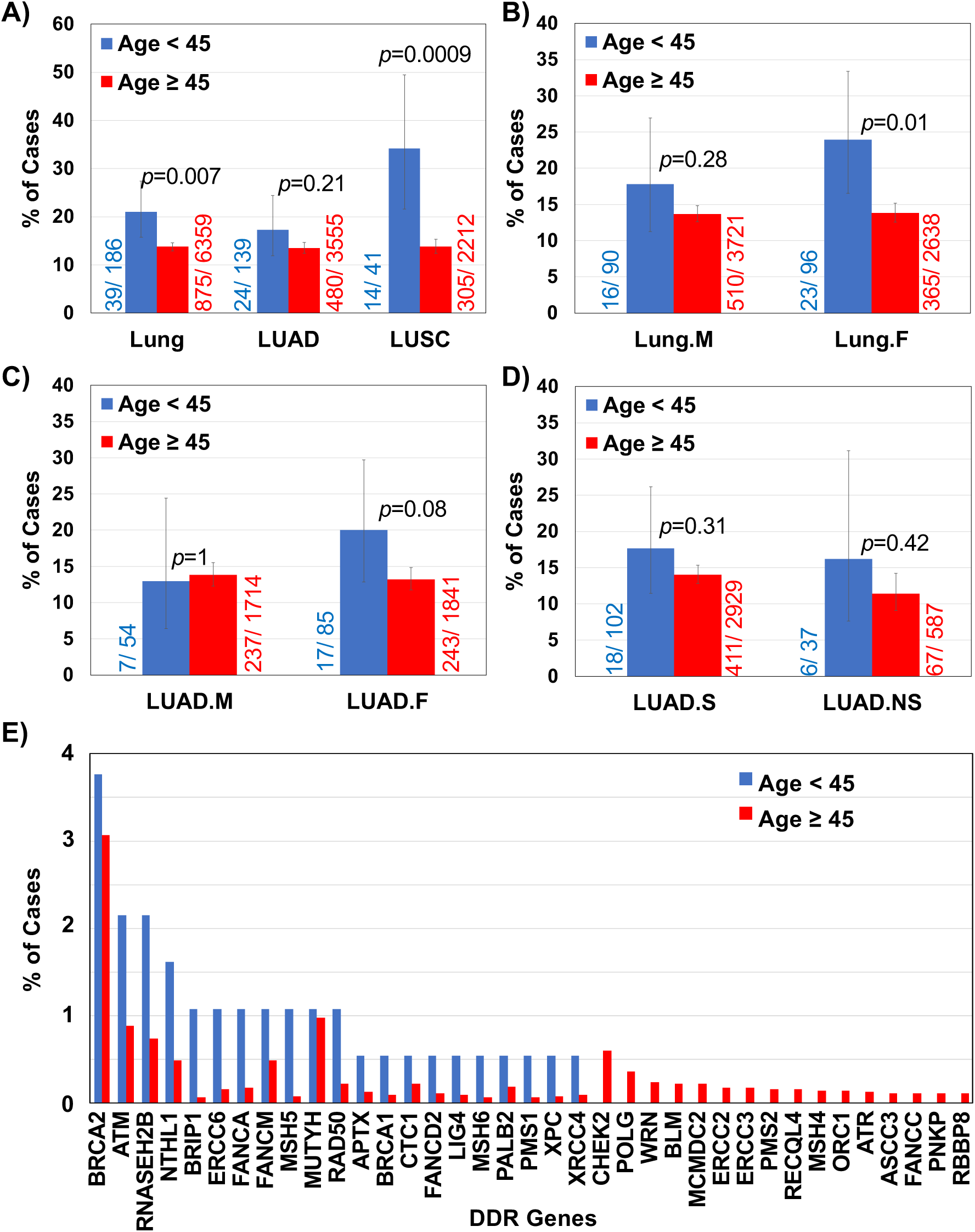
The frequency of rare germline P/LP variants (gnomAD NFE MAF < 5%) in DDR genes in young adults (< 45 years, blue) versus older (≥ 45 years, red) lung cancer patients with 95% confidence intervals. **A)** all lung cancer patients; LUAD patients; and LUSC patients; **B)** male lung cancer patients; female lung cancer patients; **C)** male LUAD patients; female LUAD patients; **D)** smoker LUAD patients; and never-smoker LUAD patients; and **E)** gene-level distribution of rare P/LP variants in DDR genes in lung cancer patients. (Listing genes observed in > 0.1% of patients).

Beyond DDR genes, young adult patients also showed enrichment in inborn errors of immunity (IEI) genes associated with antibody deficiencies, as well as several MSigDB hallmark gene sets (Figure 3, Supplementary Table S6). Across the full cohort, 914 patients (14.0%) harbored at least one rare P/LP variant in DDR genes, 727 (11.1%) in genes associated with inborn errors of immunity, 539 (8.2%) in cancer driver genes, and 369 (5.6%) in established lung cancer risk genes. Because DDR genes exhibited the strongest prevalence, subsequent analyses focused primarily on this gene set.

### Enrichment of DDR germline variants in young-onset is strongest among females

We next asked whether the enrichment of DDR variants differed by sex (Figure 4B). Young-onset patients of both sexes exhibited higher prevalence of germline DDR P/LP variants compared to older patients, with statistically significant enrichment among females (OR = 1.96; *p*-value = 0.01; 95% CI: 1.16-3.22; Figure 4B). Restricting the analysis to LUAD revealed a similar trend among females, although this did not reach statistical significance (OR = 1.64; *p*-value = 0.08; 95% CI: 0.89-2.89; Figure 4C).

### DDR germline variant enrichment in young-onset persists across smoking status in LUAD

Given the high prevalence of smoking among LUSC patients (>90%), we further examined LUAD stratified by smoking status and age group. Among never-smokers, DDR P/LP variants were more frequent in young-onset patients (16%) compared to older patients (11%; Figure 4D), although statistical power was limited. A similar pattern was observed among smokers (18% vs. 14%), suggesting that enrichment of DDR germline variants in young-onset disease is not solely explained by smoking exposure (Figure 4D).

### *BRIP1, ERCC6*, and *MSH5* contribute to enrichment of germline DDR variants in young-onset lung cancer

We next examined which individual DDR genes contributed to the observed enrichment. Genes that exhibited a higher prevalence of rare germline P/LP variants among young-onset patients include *BRCA2, ATM, RNASEH2B, NTHL1, BRIP1, ERCC6, FANCA, FANCM, MSH5, MUTYH, RAD50, APTX, BRCA1, CTC1, FANCD2, LIG4, MSH6, PALB2, PMS1, XPC* and *XRCC4* (Figure 4E). Among these, *BRIP1* (OR=17.24; *p*=0.01; 95% CI=1.55-120.80), *ERCC6* (OR= 6.90; *p*=0.04; 95% CI= 0.73-32.72), and *MSH5* (OR= 13.79; *p*=0.02; 95% CI= 1.31-84.76) reached statistical significance (*p* ≤ 0.05). However, confidence intervals were wide due to small numbers of carriers. The complete list of DDR genes with P/LP variants in the study population is provided in Supplementary Table S7.

Note that among more common DDR variants (gnomAD NFE MAF > 1%), we observed an additional *MSH5* variant, rs28399976, which also trended toward enrichment in young-onset patients (8/186 vs. 167/6,359; OR=1.67; p=0.16; 95% CI= 0.70-3.43), although individually it was not statistically significant. Most carriers were smokers in both age groups (6/8 vs. 153/167), consistent with prior reports linking *MSH5* variation with smoking-associated lung cancer^46^. However, due to conflicting pathogenicity annotations, this variant was excluded from primary analyses above.

### *Fanconi Anemia* pathway genes were enriched in young-onset lung cancer

Building on our prior work showing that germline P/LP variants in Fanconi Anemia (FA) pathway genes increase risk of LUSC^6^, we assessed whether variants across 22 FA pathway genes^6^ were enriched in young-onset patients. Consistent with the broader DDR signal, rare P/LP burden of FA pathway was higher in young-onset patients compared with older patients (OR=1.96; *p*-value=0.02; 95% CI= 1.08-3.33; Supplementary Figure S3). This enrichment was most pronounced in LUSC (OR=4.85; *p*-value=3.89e-04; 95% CI= 1.99-10.71), supporting a role for FA pathway defects in both increased risk and earlier onset of LUSC among affected individuals. Enrichment remained significant among young-onset males (OR=2.35; *p*-value=0.04; 95% CI= 1.02-4.79), with similar but non-significant trends observed among females and within LUAD across smoking strata (Supplementary Figure S3).

### Germline variants in cancer driver genes trended towards higher frequency in young-onset patients

We next compared the frequency of rare P/LP variants in cancer driver gene-set between young-onset and older patients (Supplementary Figure S5A). Similar to the pattern observed in DDR genes, young-onset patients exhibited a higher prevalence of driver gene variants across all lung cancers overall and within LUAD. This pattern remained consistent after stratification by sex and, within LUAD by smoking status (Supplementary Figure S5B-C), although these differences did not reach statistical significance.

### Secondary analyses on common variants for polygenic risk

#### Young-onset lung cancer patients exhibit higher polygenic risk scores

To complement our rare variant analyses, we assessed the contribution of common genetic variation using a validated lung cancer polygenic risk score (PRS)^45^ in the Sinai-Harvard-Yale cohort. As a non-cancer comparator, we included 500 European individuals from the 1000 Genomes Project. Young-onset patients exhibited statistically significantly higher PRS values compared with older patients (Figure 5), supporting differences in inherited genetic architecture between early- and late-onset disease. As expected, both patient groups had higher PRS values than controls, supporting the validity of the PRS model in this cohort (Figure 5).

**Figure 5.**
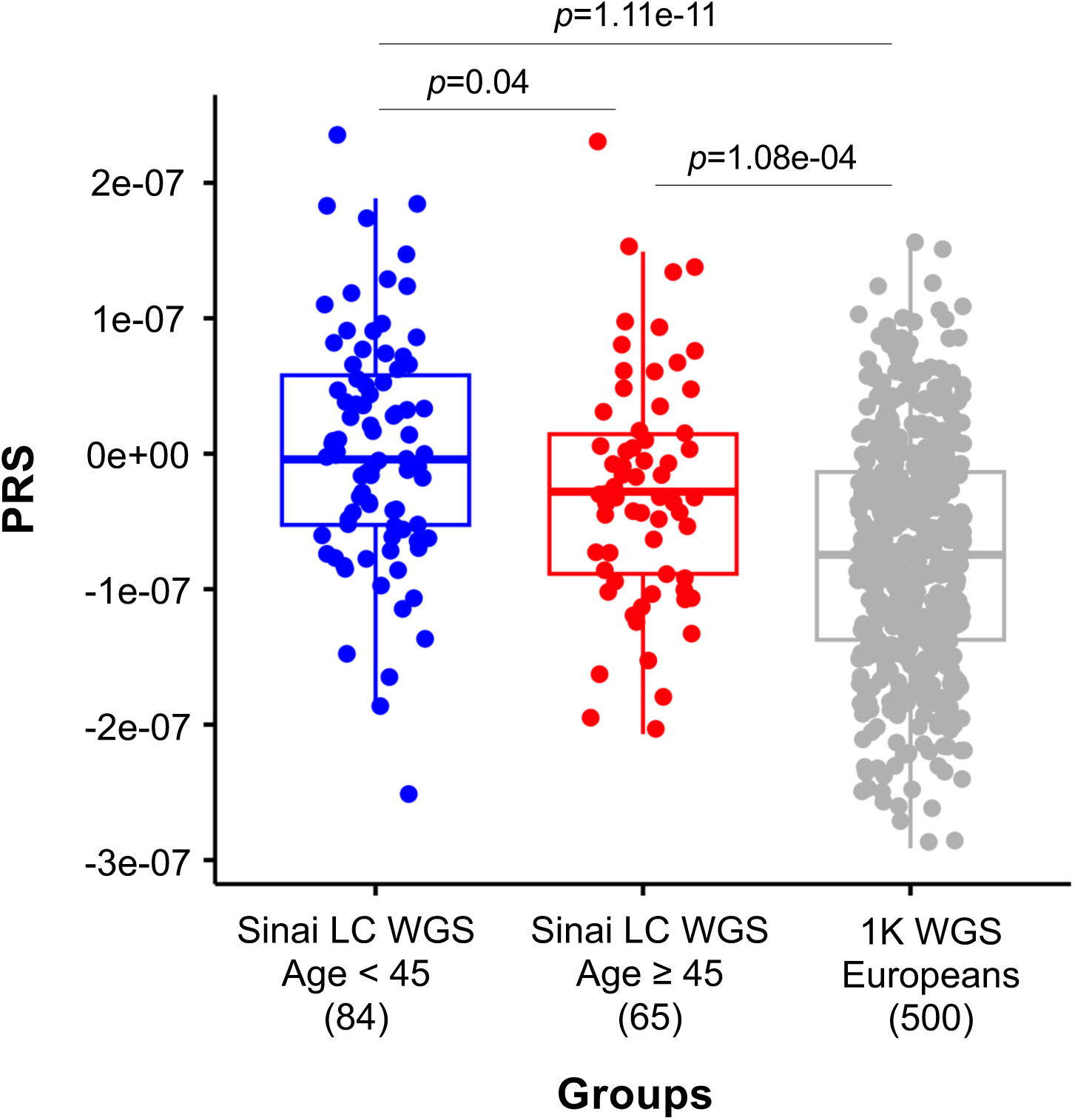
Comparison of genome-wide polygenic risk scores (PRS) among Sinai-Harvard-Yale Lung WGS patients < 45 years (n=84; blue), Sinai-Harvard-Yale Lung WGS patients ≥ 45 years (n=65; red) and 1000 Genomes WGS Europeans (n=500; gray)

### Secondary analyses on known lung cancer risk variants

#### 1. *BRCA2 rs11571833* was enriched in older lung cancer patients

Rare germline P/LP variants in *BRCA2* have been associated with increased LUSC risk in European ancestry individuals, particularly *rs11571833* (p.Lys3326Ter)^6,47^. Although classified as benign in ClinVar^27^, this variant introduces a stop codon upstream of a known pathogenic loss-of-function variant. In our cohort, rs11571833 was more prevalent in older patients, and showed a clear age-dependent gradient, with increasing prevalence across older age groups (Supplementary Figure S4), consistent with a role in later-onset disease.

#### 2. Germline *EGFR* gene variants are exceedingly rare in lung cancer patients

Given the clinical importance of EGFR in lung cancer, particularly the somatic *T790M* mutation as a main reason for EGFR-tyrosine kinase inhibitor (TKI) acquired resistance in ∼60% of tumors treated with first-generation TKIs^48^, we evaluated the prevalence of germline EGFR variants in our cohort. Although rare in the general population, germline *T790M* and other variants (*V843I, R776H* and *P848L)* have also been associated with lung cancer risk^49–51^. However, the germline *T790M* variant is not reported as P/LP in ClinVar^27^, and other *EGFR* variants are reported as either variants of uncertain significance (VUS) or with conflicting interpretations. Accordingly, they were not included in our primary analyses. Nonetheless, given their clinical relevance, we examined their prevalence and found that EGFR variants were exceedingly rare: only 2 older patients (%0.0315) harbored the *T790M* variant, and 5 other patients (1 young-onset and 4 older) carried the *P848L* variant. These findings suggest that, while the oft discussed *EGFR* germline variants are clinically important, given their rarity, there is much to be gained from understanding the broader germline genome of lung cancer patients for targeted therapies.

## Discussion

Lung cancer is common among older adults but remains relatively rare before age 45. Consequently, young-onset lung cancer has been underrepresented in genomic studies. By assembling the largest germline sequencing cohort of young-onset lung cancer to date and systematically comparing it with older patients, we show that early-onset disease is characterized by a distinct inherited genetic architecture. Specifically, young-onset patients harbor a higher burden of rare pathogenic or likely pathogenic (P/LP) germline variants in DNA damage response (DDR) genes and exhibit higher polygenic risk scores (PRSs). Together, these findings support that inherited genetic susceptibility plays a larger role in lung cancer risk in younger adults.

The most consistent signal across our analyses was the enrichment of rare germline P/LP variants in DDR genes. This pattern was observed across histology, sex, and smoking strata, and was particularly strong in lung squamous cell carcinoma (LUSC) and within the Fanconi Anemia (FA) pathway. Given the central role of DDR pathways in maintaining genomic integrity, inherited defects may lower the threshold for malignant transformation, enabling tumor development earlier in life, even in the absence of prolonged environmental exposures. These findings extend our prior work that revealed the association between rare P/LP variants in FA pathway genes and LUSC, and position germline DDR deficiency as a key mechanism underlying early-onset disease.

At the gene level, multiple DDR genes, including *BRIP1, ERCC6*, and *MSH5*, showed enrichment in young-onset patients, suggesting that germline susceptibility extends beyond a small set of historically emphasized genes. Notably, clinically recognized germline variants such as *EGFR T790M* and *TP53* alterations were exceedingly rare in our cohort. In contrast, rare P/LP germline variants across a broader spectrum of DDR genes were associated with young-onset disease. Interestingly, the *BRCA2 rs11571833* (p.Lys3326Ter) variant, previously associated with lung cancer risk^6,47^, was more prevalent in older patients and exhibited a clear age-dependent gradient, suggesting that distinct germline mechanisms may associate with lung cancer across the lifespan.

Beyond DDR, we observed enrichment of rare P/LP variants in cilia-related genes and pathways, representing a potentially novel contribution to lung cancer susceptibility. Cilia are microtubule-based, hair-like projections on the cell surface of epithelial cells that function as sensory and signaling organelles^52^. In the lung, they are critical for epithelial homeostasis, mucociliary clearance, and key signaling pathways including Hedgehog signaling, the enrichment of which we also observed in its active ‘On’ state^53,54^. Disruption of ciliary structure and function may impair airway defense, promote chronic inflammation, and increase susceptibility to infection and environmental carcinogens^55^. Complementing these, we also observed enrichment of cell-adhesion related genes, pointing to impaired epithelial integrity and altered cell-cell interactions^56^. Consistent with these findings, disease enrichment analysis linked young-onset lung cancer to chronic obstructive pulmonary disease (COPD), an established lung cancer risk factor^57^. Taken together, these results suggest that inherited defects in epithelial structure and signaling may contribute to lung cancer risk in younger adults.

In addition, we observed enrichment of rare germline P/LP variants in immune-related pathways, including the complement system and inborn errors of immunity (IEI), with the strongest signal in antibody deficiency. The lung is continuously exposed to microbial and environmental insults, and effective immune surveillance is critical for maintaining tissue homeostasis and eliminating damaged or transformed cells. The complement system is central to host defense, lung injury responses, clearance of abnormal cells, and regulation of anti-tumor immunity, and is tightly linked to humoral immunity through antibody-antigen complexes that activate the classical complement pathway^58–61^. The enrichment in both antibody deficiency and complement pathways may reflect disruption of shared humoral immune surveillance mechanisms in early lung cancer development^58^.

At the variant level, P/LP variants in core complement genes C2, C3, and C9 were more prevalent in young-onset patients, with C3 reaching statistical significance. Loss-of-function (LOF) variants in C2 (rs9332736) and C9 (rs34000044) impact distinct steps in complement activation^62,63^, potentially impairing complement function and reducing effective immune surveillance. In younger individuals, where cumulative mutagen exposure is limited, such defects may represent a predisposing factor for lung cancer^64^. Complement and antibody deficiencies are also associated with recurrent respiratory infections, chronic inflammation, and autoimmune disorders, processes that are closely linked to COPD, and may further promote carcinogenesis through persistent tissue injury and the development of a pro-tumorigenic microenvironment^65–68^. Young adults with such immune abnormalities, particularly those experiencing recurrent pulmonary infections, may therefore represent a population at elevated risk for lung cancer.

Notably, although early studies suggested increased cancer incidence among individuals with primary immunodeficiencies, more recent studies have not consistently demonstrated a strong association with solid tumors, including lung cancer^69^. However, given the rarity of young-onset lung cancer, specific immunodeficiencies, such as germline complement or antibody abnormalities may confer risk in a context-dependent manner not detectable in aggregate analyses. Together, our findings suggest a more nuanced relationship between inherited immune dysfunction and early lung cancer development that warrants focused future investigation.

Our analysis of common genetic variation further supports age-dependent differences in inherited susceptibility. Young-onset lung cancer patients exhibited significantly higher PRS values compared with older patients. Together with the enrichment of rare P/LP variants, including those in DDR and immune-related pathways, these results indicate that both rare and common inherited variants contribute to lung cancer risk in younger individuals.

These findings have potential implications for lung cancer screening. Although low-dose computed tomography (LDCT) reduces lung cancer mortality^70–72^, current screening criteria are largely based on age (50-80 years) and smoking history^73^, leaving a substantial proportion of cases, including many in younger individuals, outside eligibility^74,75^. Our results suggest that germline genetic risk factors, including certain rare P/LP variants and elevated PRS, may help identify individuals who could benefit from earlier or more intensive screening. Integrating germline genetic information into risk prediction models may therefore improve early detection strategies and enable more personalized screening approaches. These findings also support consideration of genetic counseling and cascade testing of unaffected family members who may share inherited risk. Collectively, integrating germline genetic risk into screening paradigms could enhance early detection and precision prevention strategies^76^, and may additionally support tobacco cessation efforts.

Germline DDR variants may also have therapeutic implications. In several tumor types, both germline and somatic mutations in DDR genes confer sensitivity to poly (ADP-ribose) polymerase (PARP) inhibitors and platinum-based chemotherapy^77^. A recent study further demonstrated that germline-mutated small cell lung cancers can exhibit favorable responses to DDR-targeted therapies^78^. Emerging evidence also suggests that DDR alterations may interact with immunotherapies, impacting patient selection^79–82^, and that certain DNA damaging chemotherapies may promote immunogenic tumor cell death^83^. Together, these findings support further investigation of DDR-targeted therapeutic strategies in lung cancer patients with germline DDR alterations.

There were certain methodological considerations for germline gene discovery in cancer risk. Standard rare variant discovery analysis pipelines often apply strict MAF thresholds or exclude variants with uncertain pathogenicity. However, several variants associated with lung cancer risk, including *BRCA2* rs11571833^6,47^, would be excluded under such criteria despite epidemiologic evidence of association; it is currently not listed as a P/LP in ClinVar^27^. Similarly, we observed that the *MSH5* rs28399976 variant, involved in DNA mismatch repair and previously shown to harbor common lung cancer risk variants^84^, was at MAF 1.7% in gnomAD^85^ NFE cohort, and would be filtered out in standard pipelines^86,87^ (e.g. MAF<1%) and excluded due to uncertain pathogenicity. These observations underscore that using strict filter cutoffs may limit discovery, particularly for moderately penetrant variants. In addition, many Variants of Uncertain Significance (VUS) remain uncharacterized in the context of lung cancer, highlighting the need for lung cancer–specific variant interpretation frameworks and functional studies.

This study should be considered in the context of its limitations. Although we assembled the largest germline sequencing cohort to date on young-onset lung cancer, the rarity of individual germline variants limits statistical power for single-gene analyses. Some observed trends met statistical significance thresholds without correction for multiple testing, and it is likely that additional trends we observed would be significant if we had a larger sample size. Larger studies will be necessary to confirm these findings. For computational efficiency, we did not realign and jointly call the publicly available exome data. While both young and old cases are present in each cohort, we cannot exclude differences in variant detection between cohorts with different frequencies of young cases resulting in false positives. In addition, due to the composition of the available biobank and consortium studies, our results are limited to individuals of European ancestry, reflecting the composition of available datasets and underscoring the need for studies in more diverse populations to improve generalizability and advance health equity. Finally, genotype calls were derived from study-specific pipelines rather than reprocessed through a unified framework, which may introduce variability and potential bias given age distributions and sequencing approaches across cohorts.

## Data Availability

The UK Biobank data are available on application to the UK Biobank (https://www.ukbiobank.ac.uk/). CPTAC data are available via the National Cancer Institute Proteomic Data Commons (https://pdc.cancer.gov/). EAGLE cohort data are available in dbGaP (http://www.ncbi.nlm.nih.gov/gap) through dbGaP accession number phs002496 and additional data were obtained through a data transfer agreement between Mount Sinai and National Cancer Institute. TCGA, TRICL-LUSC and TRICL-LCINIS-NYC-LUAD data used in this study were obtained from our previously published sources.

## Acknowledgements

This work was supported by grants to Z.H.G from LUNGevity Foundation, Uniting Against Lung Cancer Foundation and National Cancer Institute (NCI) Cancer Moonshot R33 grant CA263705-01; to R.J.K. from the NCI R01 grants CA167824 and CA244948; to D.C.C from National Institutes of Health (NIH) grant 2U01CA209414 supporting the Boston Lung Cancer Study (BLCS); to A.A.P from NIH grant U01 CA233364; to B.E.G.R from the NIH K08 award CA151645-01 and P50 award 196530-01 and in part through the computational resources and staff expertise provided by Scientific Computing at the Icahn School of Medicine at Mount Sinai and supported by the Clinical and Translational Science Awards (CTSA) grant UL1TR004419 from the National Center for Advancing Translational Sciences.

## Author contributions

M.E.S, R.J.K. and Z.H.G. wrote the manuscript. Z.H.G, B.E.G.R, A.A.P and D.C.C recruited patients and collected samples. M.E.S, R.J.K. and Z.H.G. performed whole genome sequencing analysis (variant calling) and other data analyses. J.S provided EAGLE cohort data. Z.H.G. conceived, designed and supervised the study. All authors approved the final manuscript.

## Notes

**Funding:** This work was supported by grants from LUNGevity Foundation, Uniting Against Lung Cancer Foundation, National Institutes of Health (NIH) and National Cancer Institute (NCI)

**Competing Financial Interests:** The authors declare no competing financial interests.

### Competing Interest Statement

The authors have declared no competing interest.

### Author Declarations

Yale samples were collected under HIC #1010007459. The Boston Lung Cancer Study cohort was approved by the Mass General Brigham (MGB) IRB, protocol #1999P004935.

