## supplemental materials for "Inherited genetic risk factors in young-onset lung cancer"

**Funding**
This work was supported by grants from LUNGevity Foundation, Uniting Against Lung Cancer Foundation, National Institutes of Health (NIH) and National Cancer Institute (NCI)

**Competing Financial Interests**
The authors declare no competing financial interests.

**Supplementary Data**

**Supplementary Table S1:** Study cohorts

**Supplementary Table S2:** Number of specific types of pathogenic/likely pathogenic (P/LP) variants

**Supplementary Table S3:** Genes with rare P/LP variants that exhibit significant difference in prevalence between younger and older lung cancer patients, A) Genes with higher prevalence in younger patients B) Genes with higher prevalence in older patients.

**Supplementary Table S4:** Over-represented gene-sets ($p$<0.01) among the 38 genes enriched for P/LP variants (Supp. Table S3) between the two age groups.

**Supplementary Table S5:** List of genes i) DDR genes ii) Cancer driver genes

**Supplementary Table S6:** Gene-sets with rare P/LP variants that show a significant difference in prevalence between younger and older lung cancer patients

**Supplementary Table S7:** DDR genes with P/LP variants in lung cancer patients i) Age < 45 years ii) Age ≥ 45 years

**Supplementary Figure S1:** Principal Component Analysis (PCA) of Sinai-Harvard-Yale Young Lung Cancer cohort.

**Supplementary Figure S2:** Prevalence of rare germline P/LP variants in DNA damage repair genes across age groups in lung cancer patients with standard error bars.

**Supplementary Figure S3:** The comparative frequency of rare germline P/LP variants in *Fanconi Anemia* genes in young adult (< 45 years, blue) versus older (≥ 45 years, red) patients.

**Supplementary Figure S4:** The frequency of germline variant rs11571833 (*BRCA2* - p.Lys3326Ter) in young adults (< 45 years, blue) versus older (≥ 45 years, red) patients with 95% confidence intervals. A) all lung cancer participants; LUAD participants only; LUSC participants only and B) male lung cancer participants; female lung cancer participants C) prevalence across age groups in lung cancer patients with standard error bars.

**Supplementary Figure S5:** The comparative frequency of rare germline P/LP variants in cancer driver genes in young adult (< 45 years, blue) versus older (≥ 45 years, red) patients.

**Supplementary Table S1: Study cohorts**

| Cohort | Median age | # LUAD samples | # LUSC samples | # Other samples |
|---|---|---|---|---|
| TCGA[a] | 68 | 410 | 434 | - |
| CPTAC[b] | 68 | 41 | 82 | - |
| TRICL-LUSC[c] | 64 | - | 318 | - |
| TRICL-LCINIS-NYC_institute LUAD[d] | 63 | 483 | - | - |
| EAGLE | 68 | 662 | 479 | 200 |
| UK Biobank | 69 | 1,964 | 926 | 397 |
| Lung WGS | 44 | 134 | 14 | 1 |

**Supplementary Table S2: Number of specific types of pathogenic/likely pathogenic (P/LP) variants.**

| Variant Type | # P/LP Variants (53,491) | # P/LP Variants with MAF < 5% (20,556) | # P/LP Variants with MAF < 1% (17,559) |
|---|---|---|---|
| frameshift deletion | 9,484 | 4,622 | 4,518 |
| frameshift insertion | 7,337 | 4,049 | 3,064 |
| frameshift substitution | 1 | 1 | 1 |
| ncRNA_exonic | 78 | 78 | 68 |
| ncRNA_exonic,splicing | 10 | 10 | 10 |
| nonframeshift deletion | 157 | 39 | 39 |
| nonframeshift insertion | 8 | 8 | 8 |
| nonframeshift substitution | 1 | 1 | 1 |
| nonsynonymous SNV | 13,541 | 5,334 | 4,470 |
| splicing | 815 | 815 | 815 |
| stopgain | 22,003 | 5,543 | 4,509 |
| stoploss | 3 | 3 | 3 |
| synonymous SNV | 32 | 32 | 32 |
| unknown | 21 | 21 | 21 |

**Supplementary Table S3: Genes with rare P/LP variants that exhibit significant difference in prevalence between younger and older lung cancer patients, A) Genes with higher prevalence in younger patients B) Genes with higher prevalence in older patients.**

**Supplementary Table S3A:**

| Gene | # Cases (%) Age < 45 | # Cases (%) Age ≥ 45 | OR | *p*-value | 95% CI: Lower | 95% CI: Upper |
|---|---|---|---|---|---|---|
| GPR161 | 10 (5.38) | 16 (0.25) | 22.48 | 9.66E-10 | 8.99 | 53.55 |
| LFNG | 6 (3.23) | 12 (0.19) | 17.60 | 6.79E-06 | 5.36 | 51.29 |
| TPK1 | 15 (8.06) | 145 (2.28) | 3.76 | 4.38E-05 | 2.01 | 6.58 |
| MYO7A | 5 (2.69) | 15 (0.24) | 11.67 | 1.92E-04 | 3.28 | 34.25 |
| HGSNAT | 3 (1.61) | 2 (0.03) | 52.00 | 2.17E-04 | 5.91 | 615.83 |
| PRKRA | 5 (2.69) | 18 (0.28) | 9.72 | 3.90E-04 | 2.79 | 27.56 |
| DNAJC30 | 3 (1.61) | 5 (0.08) | 20.79 | 1.14E-03 | 3.20 | 107.89 |
| TRIM63 | 2 (1.08) | 1 (0.02) | 68.82 | 2.37E-03 | 3.57 | 3943.34 |
| PRDM12 | 2 (1.08) | 1 (0.02) | 68.82 | 2.37E-03 | 3.57 | 3943.34 |
| C3 | 2 (1.08) | 1 (0.02) | 68.82 | 2.37E-03 | 3.57 | 3943.34 |
| FLG | 16 (8.60) | 235 (3.70) | 2.45 | 2.61E-03 | 1.35 | 4.18 |
| NEXN | 2 (1.08) | 2 (0.03) | 34.47 | 4.64E-03 | 2.49 | 472.52 |
| MLIP | 2 (1.08) | 2 (0.03) | 34.47 | 4.64E-03 | 2.49 | 472.52 |
| NRL | 2 (1.08) | 2 (0.03) | 34.47 | 4.64E-03 | 2.49 | 472.52 |
| PRF1 | 2 (1.08) | 2 (0.03) | 34.47 | 4.64E-03 | 2.49 | 472.52 |
| PHKB | 3 (1.61) | 11 (0.17) | 9.45 | 6.52E-03 | 1.68 | 36.21 |
| DNAH5 | 4 (2.15) | 23 (0.36) | 6.05 | 6.64E-03 | 1.51 | 17.96 |
| PDE11A | 8 (4.30) | 93 (1.46) | 3.03 | 7.77E-03 | 1.25 | 6.35 |
| ABCD4 | 2 (1.08) | 4 (0.06) | 17.24 | 0.01 | 1.55 | 120.80 |
| ZMYND15 | 2 (1.08) | 4 (0.06) | 17.24 | 0.01 | 1.55 | 120.80 |
| BRIP1 | 2 (1.08) | 4 (0.06) | 17.24 | 0.01 | 1.55 | 120.80 |
| GBE1 | 3 (1.61) | 15 (0.24) | 6.93 | 0.01 | 1.27 | 24.79 |
| ARMC4 | 2 (1.08) | 5 (0.08) | 13.79 | 0.02 | 1.31 | 84.76 |
| MSH5 | 2 (1.08) | 5 (0.08) | 13.79 | 0.02 | 1.31 | 84.76 |
| ACAD8 | 2 (1.08) | 5 (0.08) | 13.79 | 0.02 | 1.31 | 84.76 |
| CNTN2 | 2 (1.08) | 6 (0.09) | 11.49 | 0.02 | 1.13 | 64.84 |
| ABHD14A-ACY1 | 2 (1.08) | 6 (0.09) | 11.49 | 0.02 | 1.13 | 64.84 |
| DNAH9 | 3 (1.61) | 19 (0.30) | 5.47 | 0.02 | 1.03 | 18.81 |
| ENAM | 2 (1.08) | 7 (0.11) | 9.85 | 0.03 | 0.99 | 52.18 |
| PITRM1 | 2 (1.08) | 7 (0.11) | 9.85 | 0.03 | 0.99 | 52.18 |
| PLA2G2A | 2 (1.08) | 8 (0.13) | 8.62 | 0.03 | 0.89 | 43.59 |
| NECTIN3 | 2 (1.08) | 10 (0.16) | 6.90 | 0.04 | 0.73 | 32.72 |
| IQCE | 2 (1.08) | 10 (0.16) | 6.90 | 0.04 | 0.73 | 32.72 |
| ERCC6 | 2 (1.08) | 10 (0.16) | 6.90 | 0.04 | 0.73 | 32.72 |
| GALNS | 2 (1.08) | 10 (0.16) | 6.90 | 0.04 | 0.73 | 32.72 |
| HIF1A | 3 (1.61) | 26 (0.41) | 3.99 | 0.05 | 0.77 | 13.21 |

**Supplementary Table S3B:**

| Gene | # Cases (%) Age < 45 | # Cases (%) Age ≥ 45 | OR | *p*-value | 95% CI: Lower | 95% CI: Upper |
|---|---|---|---|---|---|---|
| HYDIN | 20 (10.75) | 1,316 (20.70) | 0.46 | 5.85E-04 | 0.27 | 0.74 |
| IVD | 20 (10.75) | 1,073 (16.87) | 0.59 | 0.03 | 0.35 | 0.95 |

**Supplementary Table S4: Over-represented gene-sets (p<0.01) among the 38 genes enriched for P/LP variants (Supp. Table S3) between the two age groups.**

### Jensen DISEASES Experimental 2025

| Term | Overlap | p-value | q-value | Odds ratio | Combined score | Genes |
|---|---|---|---|---|---|---|
| Chronic Obstructive Pulmonary Disease | 6/1113 | 0.0176 | 0.4998 | 3.19 | 12.91 | GALNS;DNAH5;MSH5; IQCE;NEXN;CNTN2 |
| Obstructive Lung Disease | 6/1113 | 0.0176 | 0.4998 | 3.19 | 12.91 | GALNS;DNAH5;MSH5; IQCE;NEXN;CNTN2 |
| Lung Adenocarcinoma | 2/119 | 0.0215 | 0.4998 | 9.42 | 36.20 | MSH5;NEXN |

### GO Cellular Component 2025

| Term | Overlap | p-value | q-value | Odds ratio | Combined score | Genes |
|---|---|---|---|---|---|---|
| Cilium (GO:0005929) | 5/402 | 9.30E-04 | 0.0633 | 7.47 | 52.12 | DNAH5;GPR161;HYDIN ;IQCE;DNAH9 |

### GO Biological Process 2025

| Term | Overlap | p-value | q-value | Odds ratio | Combined score | Genes |
|---|---|---|---|---|---|---|
| Homophilic Cell Adhesion via Plasma Membrane Adhesion Molecules (GO:0007156) | 3/59 | 1.91E-04 | 0.0363 | 30.47 | 260.89 | NEXN; CNTN2; NECTIN3 |
| Cilium Movement (GO:0003341) | 3/63 | 2.32E-04 | 0.0363 | 28.43 | 237.90 | DNAH5;HYDIN;DNAH9 |
| Dendrite Self-Avoidance (GO:0070593) | 2/14 | 3.15E-04 | 0.0363 | 92.36 | 744.61 | NEXN;CNTN2 |
| Eye Photoreceptor Cell Development (GO:0042462) | 2/17 | 4.70E-04 | 0.0405 | 73.88 | 566.18 | NRL;MYO7A |
| Cilium Movement Involved in Cell Motility (GO:0060294) | 2/20 | 6.54E-04 | 0.0451 | 61.56 | 451.39 | DNAH5;DNAH9 |
| Cilium-Dependent Cell Motility (GO:0060285) | 2/27 | 0.0012 | 0.0530 | 44.30 | 298.06 | DNAH5;DNAH9 |
| Positive Regulation of Vascular Endothelial Growth Factor Production (GO:0010575) | 2/27 | 0.0012 | 0.0530 | 44.30 | 298.06 | C3;HIF1A |
| Positive Regulation of G Protein-Coupled Receptor Signaling Pathway (GO:0045745) | 2/28 | 0.0013 | 0.0530 | 42.60 | 283.48 | C3;HIF1A |
| Regulation of Vascular Endothelial Growth Factor Production (GO:0010574) | 2/29 | 0.0014 | 0.0530 | 41.02 | 270.08 | C3;HIF1A |
| DNA Damage Checkpoint Signaling (GO:0000077) | 2/38 | 0.0024 | 0.0817 | 30.75 | 185.92 | BRIP1;ERCC6 |
| Regulation of Glycolytic Process (GO:0006110) | 2/42 | 0.0029 | 0.0905 | 27.67 | 161.82 | TRIM63;HIF1A |
| DNA Integrity Checkpoint Signaling (GO:0031570) | 2/47 | 0.0036 | 0.0983 | 24.59 | 138.35 | BRIP1;ERCC6 |

| | | | | | | |
|---|---|---|---|---|---|---|
| Fatty Acid Beta-Oxidation (GO:0006635) | 2/48 | 0.0038 | 0.0983 | 24.05 | 134.34 | ABCD4;IVD |
| Cell-Cell Adhesion via Plasma-Membrane Adhesion Molecules (GO:0098742) | 3/171 | 0.0042 | 0.0983 | 10.10 | 55.37 | NEXN; CNTN2; NECTIN3 |
| Sensory Perception (GO:0007600) | 2/52 | 0.0044 | 0.0983 | 22.12 | 120.10 | PRDM12;MYO7A |
| Nucleotide-Excision Repair (GO:0006289) | 2/53 | 0.0046 | 0.0983 | 21.69 | 116.93 | BRIP1;ERCC6 |
| Neurogenesis (GO:0022008) | 2/67 | 0.0072 | 0.1032 | 17.01 | 83.92 | PRDM12;ERCC6 |

### GO Molecular Function 2025

| Term | Overlap | p-value | q-value | Odds ratio | Combined score | Genes |
|---|---|---|---|---|---|---|
| Cell Adhesion Mediator Activity (GO:0098631) | 3/39 | 5.52E-05 | 0.0041 | 47.44 | 465.19 | NEXN;CNTN2;NECTIN3 |
| Cell-Cell Adhesion Mediator Activity (GO:0098632) | 2/49 | 0.0039 | 0.1201 | 23.54 | 130.53 | NEXN;CNTN2 |

### Reactome Pathways 2024

| Term | Overlap | p-value | q-value | Odds ratio | Combined score | Genes |
|---|---|---|---|---|---|---|
| Diseases of Metabolism | 6/264 | 9.68E-06 | 0.0018 | 14.32 | 165.33 | ABCD4;LFNG;GALNS; HGSNAT;GBE1;IVD |
| Diseases of Carbohydrate Metabolism | 3/34 | 3.64E-05 | 0.0034 | 55.11 | 563.33 | GALNS;HGSNAT;GBE1 |
| Mucopolysaccharidoses | 2/11 | 1.91E-04 | 0.0120 | 123.17 | 1054.54 | GALNS;HGSNAT |
| Branched-chain Amino Acid Catabolism | 2/21 | 7.22E-04 | 0.0341 | 58.31 | 421.84 | ACAD8;IVD |
| Glycogen Metabolism | 2/25 | 0.0010 | 0.0388 | 48.16 | 331.46 | GBE1;PHKB |
| Metabolism of Carbohydrates | 4/290 | 0.0022 | 0.0683 | 8.09 | 49.65 | GALNS;HGSNAT;GBE1 ;PHKB |
| Metabolism | 10/2181 | 0.0063 | 0.1491 | 2.93 | 14.85 | ABCD4;ACAD8;GALNS; HGSNAT;BRIP1;TPK1; GBE1;IVD;PLA2G2A; PHKB |
| Hedgehog 'On' State | 2/73 | 0.0085 | 0.1491 | 15.56 | 74.23 | GPR161;IQCE |

**Supplementary Table S5: List of genes i) DDR genes ii) Cancer driver genes**

| Gene-set | Genes |
|---|---|
| DDR genes | *ACD, ADPRS, AEN, ALKBH1, ALKBH2, ALKBH3, APEX1, APEX2, APITD1, APLF, APTX, ASCC3, ASF1A , ATAD5, ATM, ATMIN, ATR, ATRIP, ATRX, AUNIP, BABAM1, BARD1, BCAS2, BLM, BOD1L1, BRCA1, BRCA2, BRCC3, BRIP1, C11orf80, C7orf49, C9orf142, CCNB1IP1, CDA, CDC25A, CDC25B, CDC25C, CDC45, CDC5L, CDC6, CDC7, CDK12, CDKN2D, CDT1, CENPS, CENPX, CETN2, CHAF1A, CHD1L, CHEK1, CHEK2, CHTF18, CHTF8, CIB1, CLK2, CLSPN, CTC1, CUL4A, CUL4B, CUL5, DBF4, DCLRE1A, DCLRE1B, DCLRE1C, DCTPP1, DDB1, DDB2, DDIAS, DDX11, DGUOK, DHFR, DMC1, DNA2, DNPH1, DNTT, DONSON, DSCC1, DTYMK, DUT, DYNLL1, EID3, ELOF1, EME1, EME2, ENDOV, ERCC1, ERCC2, ERCC3, ERCC4, ERCC5, ERCC6, ERCC6L, ERCC6L2, ERCC8, ETAA1, EXD2, EXO1, EXO5, FAAP100, FAAP20, FAAP24, FAM111A, FAM175A, FAN1, FANCA, FANCB, FANCC, FANCD2, FANCE, FANCF, FANCG, FANCI, FANCL, FANCM, FBH1, FEN1, FIGNL1, GADD45A, GADD45G, GEN1, GMNN, GTF2H1, GTF2H2, GTF2H3, GTF2H4, GTF2H5, H2AFX, HELB, HELQ, HERC2, HFM1, HLTF, HMCES, HORMAD1, HPF1, HRAS1, HROB, HUS1, HUS1B, IDH1, IHO1, JMJD6, LIG1, LIG3, LIG4, LRR1, LRWD1, MAD2L2, MBD4, MCM10, MCM2, MCM3, MCM4, MCM5, MCM6, MCM7, MCM8, MCM9, MCMBP, MCMDC2, MDC1, MDM2, MEI4, MEIOB, MEN1, MGMT, MLH1, MLH3, MMS22L, MND1, MORF4L1, MPG, MPLKIP, MRE11A, MRNIP, MRPL40, MSH2, MSH3, MSH4, MSH5, MSH6, MTH1, MTOR, MUS81, MUTYH, NABP2, NBN, NEIL1, NEIL2, NEIL3, NHEJ1, NSMCE1, NSMCE2, NSMCE3, NSMCE4A, NTHL1, NUDT1, NUDT15, NUDT16, NUDT18, NUDT5, OBFC1, OGG1, ORC1, ORC2, ORC3, ORC4, ORC5, ORC6, PALB2, PARG, PARP1, PARP2, PARP3, PARP4, PARPBP, PAXIP1, PCLAF, PCNA, PER1, PKMYT1, PLK3, PLRG1, PMS1, PMS2, PMS2P1, PMS2P2, PMS2P3, PNKP, POLA1, POLB, POLD1, POLD2, POLD3, POLD4, POLE, POLE2, POLE3, POLE4, POLG, POLG2, POLH, POLI, POLK, POLL, POLM, POLN, POLQ, POT1, PPP4C, PRIM1, PRIM2, PRIMPOL, PRKDC, PRPF19, PSF1, PSF2, PSF3, PSMC3IP, PTEN, RAD1, RAD17, RAD18, RAD23A, RAD23B, RAD50, RAD51, RAD51AP1, RAD51AP2, RAD51B, RAD51C, RAD51D, RAD52, RAD54B, RAD54L, RAD9A, RAD9B, RADX, RBBP8, REC114, REC8, RECQL, RECQL4, RECQL5, REV1, REV3L, RFC1, RFC2, RFC3, RFC4, RFC5, RFWD3, RHNO1, RIF1, RMI1, RMI2, RNASEH2A, RNASEH2B, RNASEH2C, RNF168, RNF169, RNF212, RNF4, RNF8, RNMT, RPA1, RPA2, RPA3, RPA4, RRM1, RRM2, RRM2B, RTEL1, RTFDC1, RUVBL2, SAMHD1, SETMAR, SETX, SFR1, SHLD1, SHLD2, SHLD3, SHPRH, SLD5, SLF1, SLF2, SLFN11, SLX1A, SLX1B, SLX4, SLX4IP, SMARCA4, SMARCAL1, SMARCC1, SMC5, SMC6, SMUG1, SOD1, SOX4, SPATA22, SPIDR, SPO11, SPRTN, STK19, SWI5, SWSAP1, TDG, TDP1, TDP2, TEN1, TERC, TERF1, TERF2, TERT,* |

| | |
|---|---|
| | *TEX11, TICRR, TIMELESS, TINF2, TIP60, TIPIN, TK2, TONSL, TOP1, TOP1MT, TOP2A, TOP2B, TOP3A, TOP3B, TOPBP1, TP53, TP53BP1, TRAIP, TREX1, TREX2, TRIP13, TTK, TYMS, TZAP, UBE2A, UBE2B, UBE2N, UBE2T, UBE2V2, UIMC1, UNG, USP1, USP37, USP43, USP7, UVSSA, WDHD1, WDR48, WEE1, WRN, WRNIP1, XAB2, XPA, XPC, XRCC1, XRCC2, XRCC3, XRCC4, XRCC5, XRCC6, YWHAB, YWHAE, YWHAG, ZGRF1, ZNF451, ZRANB3, ZSWIM7* |
| Cancer driver genes | *ABL1, ACVR1, ACVR1B, ACVR2A, AJUBA, AKT1, ALB, ALK, AMER1, APC, APOB, AR, ARAF, ARHGAP35, ARID1A, ARID2, ARID5B, ASXL1, ASXL2, ATF7IP, ATM, ATR, ATRX, ATXN3, AXIN1, AXIN2, B2M, BAP1, BCL2, BCL2L11, BCOR, BRAF, BRCA1, BRCA2, BRD7, BTG2, CACNA1A, CARD11, CASP8, CBFB, CBWD3, CCND1, CD70, CD79B, CDH1, CDK12, CDK4, CDKN1A, CDKN1B, CDKN2A, CDKN2C, CEBPA, CHD3, CHD4, CHD8, CHEK2, CIC, CNBD1, COL5A1, CREB3L3, CREBBP, CSDE1, CTCF, CTNNB1, CTNND1, CUL1, CUL3, CYLD, CYSLTR2, DACH1, DAZAP1, DDX3X, DHX9, DIAPH2, DICER1, DMD, DNMT3A, EEF1A1, EEF2, EGFR, EGR3, EIF1AX, ELF3, EP300, EPAS1, EPHA2, EPHA3, ERBB2, ERBB3, ERBB4, ERCC2, ESR1, EZH2, FAM46D, FAT1, FBXW7, FGFR1, FGFR2, FGFR3, FLNA, FLT3, FOXA1, FOXA2, FOXQ1, FUBP1, GABRA6, GATA3, GNA11, GNA13, GNAQ, GNAS, GPS2, GRIN2D, GTF2I, H3F3A, H3F3C, HGF, HIST1H1C, HIST1H1E, HLA-A, HLA-B, HRAS, HUWE1, IDH1, IDH2, IL6ST, IL7R, INPPL1, IRF2, IRF6, JAK1, JAK2, JAK3, KANSL1, KDM5C, KDM6A, KEAP1, KEL, KIF1A, KIT, KLF5, KMT2A, KMT2B, KMT2C, KMT2D, KRAS, KRT222, LATS1, LATS2, LEMD2, LZTR1, MACF1, MAP2K1, MAP2K4, MAP3K1, MAP3K4, MAPK1, MAX, MECOM, MED12, MEN1, MET, MGA, MGMT, MLH1, MSH2, MSH3, MSH6, MTOR, MUC6, MYC, MYCN, MYD88, MYH9, NCOR1, NF1, NF2, NFE2L2, NIPBL, NOTCH1, NOTCH2, NPM1, NRAS, NSD1, NUP133, NUP93, PAX5, PBRM1, PCBP1, PDGFRA, PDS5B, PGR, PHF6, PIK3CA, PIK3CB, PIK3CG, PIK3R1, PIK3R2, PIM1, PLCB4, PLCG1, PLXNB2, PMS1, PMS2, POLE, POLQ, POLRMT, PPM1D, PPP2R1A, PPP6C, PRKAR1A, PSIP1, PTCH1, PTEN, PTMA, PTPDC1, PTPN11, PTPRC, PTPRD, RAC1, RAD21, RAF1, RARA, RASA1, RB1, RBM10, RET, RFC1, RHEB, RHOA, RHOB, RIT1, RNF111, RNF43, RPL22, RPL5, RPS6KA3, RQCD1, RRAS2, RUNX1, RXRA, SCAF4, SETBP1, SETD2, SF1, SF3B1, SIN3A, SMAD2, SMAD4, SMARCA1, SMARCA4, SMARCB1, SMC1A, SMC3, SOS1, SOX17, SOX9, SPOP, SPTA1, SPTAN1, SRSF2, STAG2, STK11, TAF1, TBL1XR1, TBX3, TCEB1, TCF12, TCF7L2, TET2, TGFBR2, TGIF1, THRAP3, TLR4, TMSB4X, TNFAIP3, TP53, TRAF3, TSC1, TSC2, TXNIP, U2AF1, UNCX, USP9X, VHL, WHSC1, WT1, XPO1, ZBTB20, ZBTB7B, ZC3H12A, ZCCHC12, ZFHX3, ZFP36L1, ZFP36L2, ZMYM2, ZMYM3, ZNF133, ZNF750* |

**Supplementary Table S6: Gene-sets with rare P/LP variants that show a significant difference in prevalence between younger and older lung cancer patients**

| Gene-sets | Age < 45 | | Age ≥ 45 | | OR | p-value | 95% CI: Lower | 95% CI: Upper |
|---|---|---|---|---|---|---|---|---|
| | % Cases | Genes w P/LP | % Cases | Genes w P/LP | | | | |
| HALLMARK_NOTCH_SIGNALING | 3.23 | LFNG | 0.20 | NOTCH3 LFNG | 16.24 | 9.69E-06 | 5.00 | 46.45 |
| IEI - ANTIBODY DEFICIENCY | 2.69 | INO80 MSH6 PIK3CG TNFRSF13B TRNT1 | 0.50 | FNIP1 MOGS MSH6 PIK3CG PIK3R1 PIK3R1 SLC39A7 TNFRSF13B TRNT1 | 5.46 | 3.66E-03 | 1.64 | 14.34 |
| HALLMARK_ALLOGRAFT_REJECTION | 4.84 | C2 PRF1 RARS1 ETS1 HIF1A BRCA1 | 1.73 | IL12B CD3E CD3G C2 ITGB2 TLR1 CD247 ZAP70 IFNGR1 CD40LG GLMN PRF1 IL12RB1 CDKN2A IL2RA IFNAR2 KRT1 CCR2 MMP9 EGFR IL2RG ITK F2 PRKCG MTIF2 HIF1A TAP1 DARS1 NCF4 TAP2 BRCA1 MRPL3 TAPBP | 2.89 | 6.55E-3 | 1.27 | 5.81 |
| DNA DAMAGE RESPONSE (DDR) | 20.97 | CTC1 APTX MUTYH NTHL1 XRCC4 LIG4 ERCC6 XPC FANCA FANCD2 FANCM BRIP1 BRCA1 BRCA2 PALB2 RAD50 MSH5 MSH6 PMS1 RNASEH2B ATM | 13.76 | ACD CTC1 DCLRE1B EXO1 POT1 ASCC3 APTX LIG3 MBD4 MUTYH NTHL1 PNKP XRCC1 DCLRE1C ERCC6L2 RNF168 XRCC4 LIG4 DDB2 ERCC2 ERCC3 ERCC4 ERCC6 ERCC8 GTF2H5 UVSSA XPA XPC FAN1 FANCA FANCB FANCC FANCD2 FANCF FANCG FANCI FANCL FANCM MCM8 MCM9 TRAIP BRIP1 BRCA1 BRCA2 PALB2 BARD1 BLM DNA2 HFM1 NBN NSMCE2 NSMCE3 RAD50 RAD51C RBBP8 SLF2 SPIDR TONSL MLH1 MLH3 MSH3 MSH4 MSH5 MSH6 PMS1 PMS2 TOP3A ATR CHEK1 DDX11 POLH RAD9B RECQL4 REV3L RNASEH2B SAMHD1 SMARCAL1 SPRTN TIMELESS WRN MCMDC2 PSMC3IP REC8 DGUOK RRM2B TK2 DTYMK ATM CHEK2 TP53 ATRX MPLKIP TREX1 SMARCA4 IDH1 HERC2 CDC45 CDT1 DONSON MCM10 MCM7 ORC1 ORC4 ORC6 POLE POLG POLG2 MTOR SETX MRE11 | 1.66 | 7.18E-03 | 1.13 | 2.40 |
| HALLMARK_P53_PATHWAY | 3.76 | XPC AK1 COQ8A CEBPA ELP1 EPHA2 EPS8L2 | 1.16 | TAP1 DDB2 FDXR PIDD1 CDKN2A PPM1D SLC19A2 TP53 XPC ADA RB1 POLH FUCA1 TP63 COQ8A RAD51C ITGB4 HINT1 ELP1 TCN2 EPHA2 SLC35D1 EPS8L2 ST14 TCHH ACVR1B | 3.32 | 7.90E-03 | 1.27 | 7.33 |
| HALLMARK_COMPLEMENT | 6.45 | C2 MMP13 F7 SERPINA1 C3 CTSC CD36 C9 COL4A2 GATA3 PIK3CG | 2.91 | C2 C1S CFB C1R F5 MMP13 F7 ADAM9 C1QC KLKB1 CR2 F10 SERPINA1 CD40LG GP9 C3 F2 CTSC CFH CD36 PDGFB CASP10 TMPRSS6 KYNU GP1BA ZFPM2 SH2B3 C9 PLA2G4A USP8 CD55 | 2.30 | 0.013 | 1.15 | 4.22 |

| | | | | | | | | |
|---|---|---|---|---|---|---|---|---|
| | | | | PLA2G7 COL4A2 SERPINC1 GATA3 CP LIPA F8 HNF4A PIK3CG | | | | |
| HALLMARK_KRAS_SIGNALING_DN | 9.14 | LFNG GAMT RYR1 STAG3 SLC12A3 ABCB11 PRKN MSH5 TEX15 COQ8A IDUA | 4.99 | FGFR3 PDE6B SLC6A3 TGM1 RYR2 AMBN LFNG ALOX12B ATP6V1B1 GAMT RYR1 SLC29A3 C5 EDAR STAG3 GP1BA MYH7 PROP1 SLC12A3 KRT1 MTHFR ABCB11 CELSR2 CHRNG CD40LG PRKN MEFV EGF MSH5 TEX15 CYP11B2 CLDN16 HSD11B2 COQ8A HNF1A TG IDUA TNNI3 NPHS1 TSHB GRID2 IRS4 IL12B ACTC1 BARD1 MYO15A SLC5A5 KRT5 | 1.92 | 0.017 | 1.08 | 3.21 |

**Supplementary Table S7: DDR genes with P/LP variants in lung cancer patients i) Age < 45 years ii) Age ≥ 45 years**

| Age < 45 years | | | Age ≥ 45 years | | |
|---|---|---|---|---|---|
| **Gene** | **# Cases** | **% Cases** | **Gene** | **# Cases** | **% Cases** |
| BRCA2 | 7 | 3.76 | BRCA2 | 195 | 3.07 |
| ATM | 4 | 2.15 | ATM | 56 | 0.88 |
| RNASEH2B | 4 | 2.15 | RNASEH2B | 47 | 0.74 |
| NTHL1 | 3 | 1.61 | NTHL1 | 31 | 0.49 |
| BRIP1 | 2 | 1.08 | BRIP1 | 4 | 0.06 |
| ERCC6 | 2 | 1.08 | ERCC6 | 10 | 0.16 |
| FANCA | 2 | 1.08 | FANCA | 11 | 0.17 |
| FANCM | 2 | 1.08 | FANCM | 31 | 0.49 |
| MSH5 | 2 | 1.08 | MSH5 | 5 | 0.08 |
| MUTYH | 2 | 1.08 | MUTYH | 62 | 0.97 |
| RAD50 | 2 | 1.08 | RAD50 | 14 | 0.22 |
| APTX | 1 | 0.54 | APTX | 8 | 0.13 |
| BRCA1 | 1 | 0.54 | BRCA1 | 6 | 0.09 |
| CTC1 | 1 | 0.54 | CTC1 | 14 | 0.22 |
| FANCD2 | 1 | 0.54 | FANCD2 | 7 | 0.11 |
| LIG4 | 1 | 0.54 | LIG4 | 6 | 0.09 |
| MSH6 | 1 | 0.54 | MSH6 | 4 | 0.06 |
| PALB2 | 1 | 0.54 | PALB2 | 12 | 0.19 |
| PMS1 | 1 | 0.54 | PMS1 | 4 | 0.06 |
| XPC | 1 | 0.54 | XPC | 5 | 0.08 |
| XRCC4 | 1 | 0.54 | XRCC4 | 6 | 0.09 |
| | | | CHEK2 | 38 | 0.60 |
| | | | POLG | 23 | 0.36 |
| | | | WRN | 15 | 0.24 |
| | | | BLM | 14 | 0.22 |
| | | | MCMDC2 | 14 | 0.22 |
| | | | ERCC2 | 11 | 0.17 |
| | | | ERCC3 | 11 | 0.17 |
| | | | PMS2 | 10 | 0.16 |
| | | | RECQL4 | 10 | 0.16 |
| | | | MSH4 | 9 | 0.14 |
| | | | ORC1 | 9 | 0.14 |
| | | | ATR | 8 | 0.13 |
| | | | ASCC3 | 7 | 0.11 |
| | | | FANCC | 7 | 0.11 |
| | | | PNKP | 7 | 0.11 |
| | | | RBBP8 | 7 | 0.11 |
| | | | FANCI | 6 | 0.09 |
| | | | CDT1 | 5 | 0.08 |
| | | | DDX11 | 5 | 0.08 |
| | | | DONSON | 5 | 0.08 |
| | | | FAN1 | 5 | 0.08 |
| | | | LIG3 | 5 | 0.08 |
| | | | MLH1 | 5 | 0.08 |
| | | | SAMHD1 | 5 | 0.08 |

| | | | | | |
|---|---|---|---|---|---|
| | | | SMARCAL1 | 5 | 0.08 |
| | | | TIMELESS | 5 | 0.08 |
| | | | XPA | 5 | 0.08 |
| | | | DCLRE1C | 4 | 0.06 |
| | | | DDB2 | 4 | 0.06 |
| | | | DGUOK | 4 | 0.06 |
| | | | DNA2 | 4 | 0.06 |
| | | | ERCC6L2 | 4 | 0.06 |
| | | | MSH3 | 4 | 0.06 |
| | | | NSMCE2 | 4 | 0.06 |
| | | | POLE | 4 | 0.06 |
| | | | POLH | 4 | 0.06 |
| | | | POT1 | 4 | 0.06 |
| | | | BARD1 | 3 | 0.05 |
| | | | ERCC8 | 3 | 0.05 |
| | | | FANCG | 3 | 0.05 |
| | | | FANCL | 3 | 0.05 |
| | | | HFM1 | 3 | 0.05 |
| | | | IDH1 | 3 | 0.05 |
| | | | MBD4 | 3 | 0.05 |
| | | | MCM8 | 3 | 0.05 |
| | | | MLH3 | 3 | 0.05 |
| | | | MPLKIP | 3 | 0.05 |
| | | | NBN | 3 | 0.05 |
| | | | REC8 | 3 | 0.05 |
| | | | TP53 | 3 | 0.05 |
| | | | DCLRE1B | 2 | 0.03 |
| | | | ERCC4 | 2 | 0.03 |
| | | | EXO1 | 2 | 0.03 |
| | | | FANCF | 2 | 0.03 |
| | | | MCM10 | 2 | 0.03 |
| | | | MCM7 | 2 | 0.03 |
| | | | MRE11 | 2 | 0.03 |
| | | | NSMCE3 | 2 | 0.03 |
| | | | ORC4 | 2 | 0.03 |
| | | | POLG2 | 2 | 0.03 |
| | | | RAD51C | 2 | 0.03 |
| | | | RAD9B | 2 | 0.03 |
| | | | REV3L | 2 | 0.03 |
| | | | RNF168 | 2 | 0.03 |
| | | | SETX | 2 | 0.03 |
| | | | SPIDR | 2 | 0.03 |
| | | | SPRTN | 2 | 0.03 |
| | | | TK2 | 2 | 0.03 |
| | | | TONSL | 2 | 0.03 |
| | | | TOP3A | 2 | 0.03 |
| | | | TRAIP | 2 | 0.03 |
| | | | TREX1 | 2 | 0.03 |
| | | | XRCC1 | 2 | 0.03 |
| | | | ACD | 1 | 0.02 |
| | | | ATRX | 1 | 0.02 |

| | | | | | |
|---|---|---|---|---|---|
| | | | CDC45 | 1 | 0.02 |
| | | | CHEK1 | 1 | 0.02 |
| | | | DTYMK | 1 | 0.02 |
| | | | FANCB | 1 | 0.02 |
| | | | GTF2H5 | 1 | 0.02 |
| | | | HERC2 | 1 | 0.02 |
| | | | MCM9 | 1 | 0.02 |
| | | | MTOR | 1 | 0.02 |
| | | | ORC6 | 1 | 0.02 |
| | | | PSMC3IP | 1 | 0.02 |
| | | | RRM2B | 1 | 0.02 |
| | | | SLF2 | 1 | 0.02 |
| | | | SMARCA4 | 1 | 0.02 |
| | | | UVSSA | 1 | 0.02 |

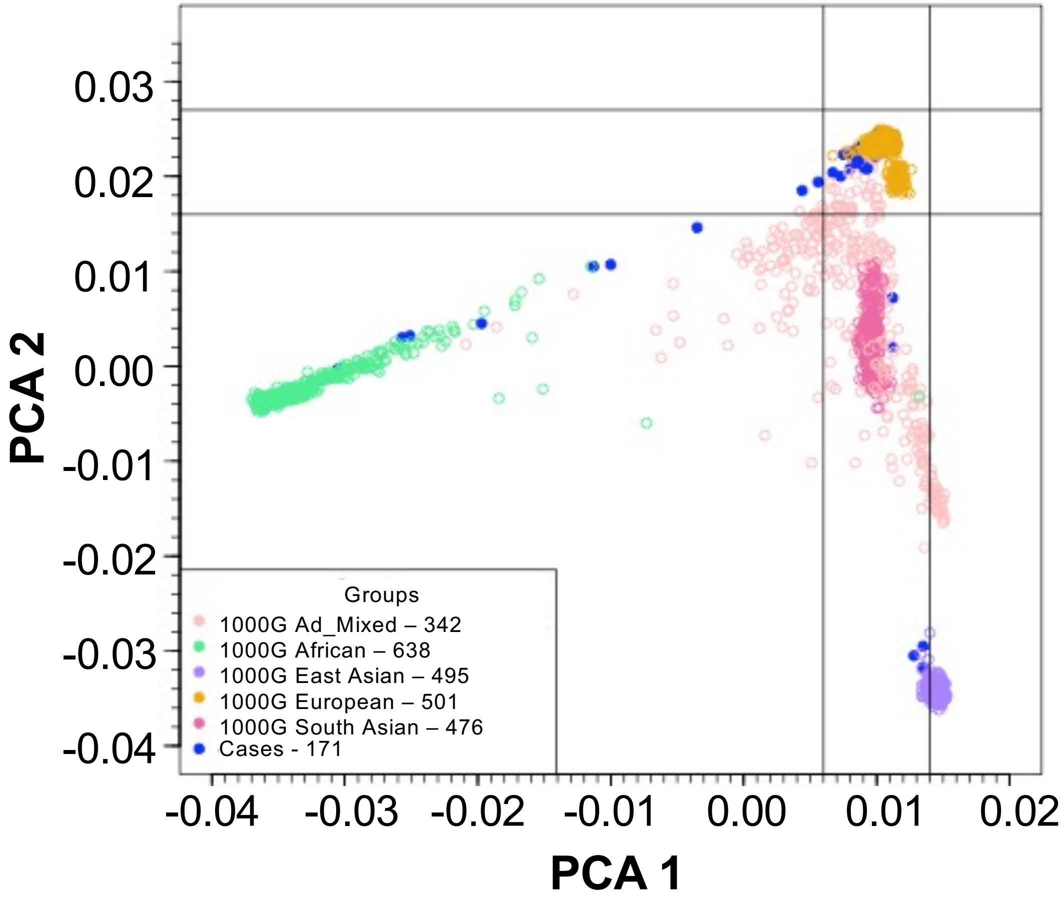


**Supplementary Figure S1: Principal Component Analysis (PCA) of Sinai-Harvard-Yale Young lung WGS cohort.** PCA based on common SNPs (MAF > 0.05) showing the top two principal components of the study cohorts together with 1000 Genomes.

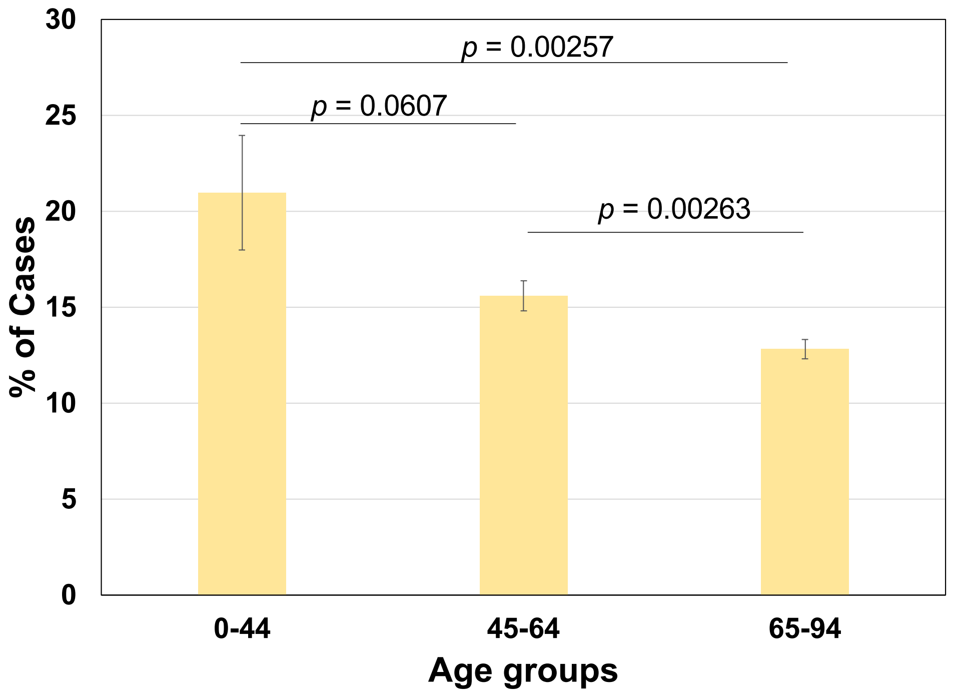


**Supplementary Figure S2:** Prevalence of rare germline P/LP variants in DNA damage repair genes across age groups in lung cancer patients with standard error bars.

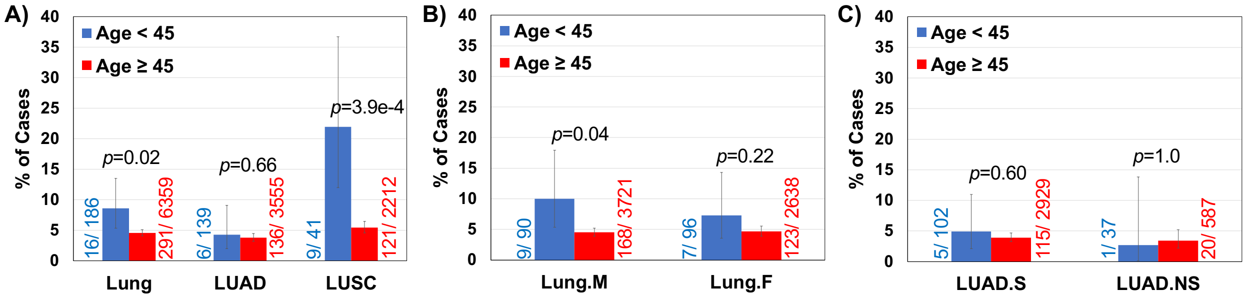


**Supplementary Figure S3: The comparative frequency of rare germline P/LP variants in *Fanconi Anemia* genes in young adult (< 45 years, blue) versus older (≥ 45 years, red)**; **A)** all lung cancer patients; LUAD patients; and LUSC patients; **B)** male lung cancer patients; female lung cancer patients; and **C)** smoker LUAD patients; and never-smoker LUAD patients.

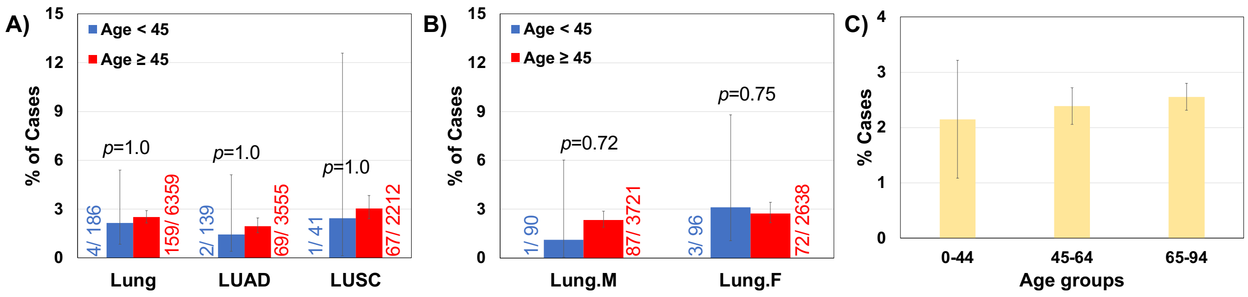


**Supplementary Figure S4: The frequency of germline variant rs11571833 (*BRCA2* - p.Lys3326Ter) in young adults (< 45 years, blue) versus older (≥ 45 years, red) patients with 95% confidence intervals. A)** all lung cancer participants; LUAD participants only; LUSC participants only and **B)** male lung cancer participants; female lung cancer participants. **C)** prevalence across age groups in lung cancer patients with standard error bars.

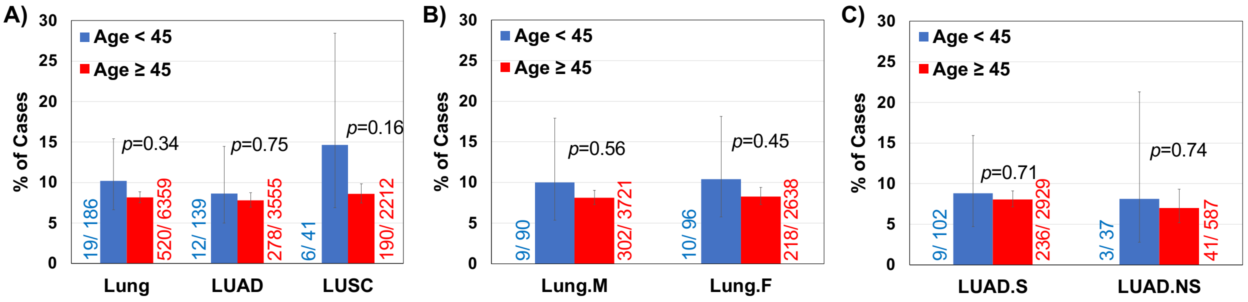


**Supplementary Figure S5: The comparative frequency of rare germline P/LP variants in cancer driver genes in young adult (< 45 years, blue) versus older (≥ 45 years, red)**; **A)** all lung cancer patients; LUAD patients; and LUSC patients; **B)** male lung cancer patients; female lung cancer patients; and **C)** smoker LUAD patients; and never-smoker LUAD patients.
